# Altered Speech Processing in Childhood Listening Difficulties as Revealed by Chirped Speech Event-Related Potentials

**DOI:** 10.64898/2026.08.13.26360392

**Authors:** Lauren Petley, Taylor R. Wicks, Lee Miller, Chelsea Blankenship, Jordan Chatwin, Brett M. Bormann, Richard S. Whittle, David R. Moore

## Abstract

**Objective:** Impaired understanding of noisy or degraded speech is a central feature of listening difficulties (LiD), but the possible causes of these symptoms are wide-ranging. Accordingly, recent research underscores the need to study these deficits using a test battery approach. Event-related potentials are useful objective metrics for studying LiD, but probing function across the speech processing hierarchy using traditional protocols is sequential and unrealistic in clinical settings. The novel chirped speech (Cheech) method combines natural speech with acoustic chirps to overcome these limitations. This study examines its utility for profiling childhood LiD.

**Methods:** Twenty-eight children (15 typically developing, 13 with LiD), aged 8-17 years old, listened to a 17-minute Cheech story and detected a target word within the story via button press while EEG data were collected from 53 scalp sites.

**Results:** Cheech successfully evoked responses from the auditory brainstem response through to the brain’s language centers, as reflected by the N400 effect. Unlike TD children, those with LiD demonstrated N400 effects with atypical distributions that favored frontal rather than the typical parietal sites. A trend towards a delayed and reduced amplitude Wave V was also observed.

**Conclusions:** Hierarchical examination of speech processing using Cheech primarily implicates altered language processing as a contributing factor to LiD, with the frontal topography of the N400 effect for those with LiD potentially suggesting a greater reliance on deliberate memory retrieval during the speech perception task.

**Significance:** LiD could arise due to auditory and/or cognitive factors. The present results demonstrate the feasibility of objective, parallel measurement across this hierarchy and point to impaired language processing as a possible mechanism.

## 1. Introduction

Approximately 5-7% of children report auditory challenges that cannot be attributed to peripheral losses, sometimes leading to a diagnosis of auditory processing disorder (Hind et al., 2011; Musiek et al., 1990). Typically recognized around 6-11 years of age (Moore et al., 2018), the disorder commonly involves difficulty understanding speech in noise, as well as rapid or degraded speech (Jerger & Musiek, 2000). In some cases, pediatric auditory processing disorder (APD) may have a very clear cause, such as a lesion or tumor of the central auditory nervous system, cerebrovascular disorders, metabolic disorders, or epilepsy (Moore, 2006). In others, its cause remains enigmatic. Inconsistencies in its diagnosis (Wilson & Arnott, 2013), as well as disagreements regarding its underlying pathology (Dillon & Cameron, 2021), have led many to prefer the term “listening difficulties” (LiD), which is based on reports of symptom presence rather than objective testing.

Due to a lack of clarity regarding APD’s underlying pathology, current diagnostic recommendations from the American Speech-Language-Hearing Association include an assortment of tests spanning auditory discrimination, auditory temporal processing and patterning, dichotic speech, monaural low-redundancy speech, and binaural interaction (American Speech-Language-Hearing Association, 2005). Beyond such clinical recommendations, research has identified a diverse array of metrics that differentiate children with LiD from their typically developing (TD) peers, including performance on nonverbal and verbal auditory tasks of varied design interrogating capacities like rhythm perception or speech perception in noise, cognitive tasks assessing abilities like working memory or attention, and language tasks evaluating skills such as vocabulary or grammar use (Falcone et al., 2026). Audiological tools have recently emerged for assessing language skills as candidate factors in LiD (Dillon et al., 2025; Zhou et al., 2026). Such tools are predicated on the well-established benefits that context provide in the comprehension of speech in noise (G. A. Miller et al., 1951). This multiplicity of options for examining LiD behaviorally is mirrored by the variety of objective measurements that have been recommended for diagnosing APD.

Event-related potentials (ERPs), including the auditory brainstem response (ABR), middle latency response (MLR), auditory steady state responses, frequency-following responses, the long-latency auditory evoked response (LLAER), P300, and the mismatch negativity have been recommended as possible APD markers for decades (Jerger & Musiek, 2000). Given the lack of evidence for the superiority of any ERP over another in the diagnostic process, and the amount of time that is needed to evoke them using traditional protocols, it is difficult to imagine how these measures could be comprehensively assessed in a single clinical visit. The present report explores the usefulness of a relatively novel method called chirped speech (“Cheech”; Miller & Moore IV, 2020). Cheech combines acoustic chirps with natural speech to permit concurrent measurements of auditory and cognitive ERPs that would otherwise require independent protocols. Its ability to evoke responses from the brainstem through to auditory cortex (i.e., from the ABR to the LLAER) has been demonstrated in healthy adults (Backer et al., 2019; Mankel et al., in press; Shehabi et al., 2025). LLAERs have also been demonstrated for children with cochlear implants (Corina et al., 2022). All of these responses have relevance in the study of LiD.

The ABR is the earliest cluster of ERPs observed in response to the onset of a sound. It is typically studied using simple sounds like tones, clicks, or chirps and comprises five to seven vertex-positive peaks (waves I –VII) that occur within 10 ms from stimulus onset (Picton, 2010). While each wave reflects contributions from several sources, the source generators of these peaks are generally thought to be the cochlea, cochlear nerve, superior olivary complex, lateral lemniscus, and inferior colliculus (Biacabe et al., 2001; Legatt et al., 1988; Melcher & Kiang, 1996). Wave V is the largest and most commonly used response, and is generated by the lateral lemniscus and inferior colliculus (Biacabe et al., 2001; Melcher & Kiang, 1996). The ABR is used clinically for neonatal hearing screening and the diagnosis of hearing loss (Joint Committee on Infant Hearing, 2007) and it is helpful in the diagnosis of several auditory and non-auditory conditions such as auditory neuropathies, Bell’s palsy, and vestibular schwannomas (Eggermont, 2019). However, its value in LiD is less clear. There is some evidence that children with suspected or diagnosed APD can show atypical amplitudes and/or latencies for some of its peaks (Allen & Allan, 2014; Gopal et al., 2002; Hunter et al., 2023; Jirsa, 2001; Omidvar et al., 2023), but such findings are not unanimous (Ankmnal-Veeranna et al., 2019; Filippini & Schochat, 2009).

The ABR is also studied using speech stimuli rather than tones, clicks, or chirps. When the stimuli are consonant-vowel syllables, it is referred to as the speech-evoked ABR (cABR) or frequency-following response (FFR; Johnson et al., 2005). Such responses consist of a transient onset response and a sustained FFR. The transient component of the speech-evoked ABR includes Waves III and V and, in children, the latency of the speech-evoked Wave V correlates modestly with that evoked by click stimuli (Song et al., 2006). Studies using speech ABRs have revealed poorer encoding of the fundamental frequency and second harmonic of speech in children who have more difficulty understanding speech in noise (Anderson et al., 2010), as well as apparent deficits in the encoding of speech timing cues for children with LiD (Filippini & Schochat, 2009; Omidvar et al., 2023; Rocha-Muniz et al., 2012).

The MLR occurs 10 - 50 ms from stimulus onset. It comprises a series of positive and negative peaks known as Na, Pa, Nb, and Pb (Picton, 2010). Like the ABR, the MLR is commonly evoked using simple sounds like tones or clicks. While the first of these peaks (Na) might originate in the inferior colliculus, the majority of the response is thought to be generated along the thalamocortical pathway, particularly in auditory cortex (Musiek & Nagle, 2018). Reduced peak amplitudes and/or prolonged latencies for the MLR have been observed in children with LiD (Abdollahi et al., 2019; Mattsson et al., 2019; Schochat et al., 2010). The MLR is followed by several later components, including P1, N1, P2, and N2, which will be referred to collectively as the LLAER. Like the ABR and MLR, the P1-N1-P2 complex largely reflects stimulus properties and occurs in response to the onset of any sound, though it can be modulated by attention (Joos et al., 2014). The neural generators for P1 are thought to include primary and secondary auditory cortices (Grunwald et al., 2003). By contrast, N1 has several subcomponents, which appear to be generated on the supratemporal plane, in auditory association cortex, and in the motor and premotor cortices (Näätänen & Picton, 1987). While P2 has some similar properties to N1, such as decreasing latencies with increased stimulus intensity and increased amplitudes at slower interstimulus intervals. Unlike N1, P2 appears to be a relatively similar response across sensory modalities (Crowley & Colrain, 2004). The N2 is linked to the interpretation of sensory stimuli, and several overlapping subcomponents of the response reflect cognitive operations (Picton, 2010). The LLAER to speech stimuli like syllables differs from that evoked by non-phonetic stimuli, particularly with respect to N1, which is attenuated and difficult to verify in syllable-evoked responses (Čeponienė et al., 2005). When evoked using speech, the N2 might reflect the encoding of higher-order auditory representations that support phoneme recognition (Čeponienė et al., 2008).

Some research has yielded evidence of LLAER alterations in children with LiD. Various latency delays have been reported for each of its peaks in APD, though the evidence is inconsistent with respect to which components are affected (Jirsa, 1992; Jirsa & Clontz, 1990; Koravand et al., 2017; Morlet et al., 2019; Tomlin & Rance, 2016). Some researchers also report reduced amplitudes for the P1-N1 complex (Tomlin & Rance, 2016) and N2 (Koravand et al., 2017), or abnormal morphologies for the peaks of the LLAER (Liasis et al., 2003). However, some studies have yielded no link between LiD and the LLAER (Mattsson et al., 2019). When evoked as part of the acoustic change complex in response to changes in the modulation rate of amplitude-modulated white noise, N1 is not significantly different between children with LiD and their TD peers (Petley et al., 2024).

Event-related potentials with longer latencies than the LLAER index cognitive operations. Depending on the task design that is used, cognitive ERPs such as P300 and N400 should also be observable with Cheech, but the present study is the first to attempt such measurements. Most notable among these in the context of audiometry is the P300, which reflects target stimulus detection and, as a general rule, is not observed in response to unattended stimuli (Duncan et al., 2009; Polich, 2007). Its functional significance is tied to selective attention and working memory updating and, accordingly, its source generators include a variety of regions in frontal and parietal cortices (Polich, 2007). Delayed and/or smaller P300 responses have been observed in children and adults with LiD with simple, target tone-detection tasks (Alonso & Schochat, 2009; Jirsa, 1992; Jirsa & Clontz, 1990; Krishnamurti, 2001; Mattsson et al., 2019). Dichotic listening with a noise masker in one ear may augment P300 latency delays for people with APD (Krishnamurti, 2001). However, like other ERP components, findings using the P300 are not uniform, and can vary depending on how measurements in this latency range are derived (Petley et al., 2024).

Unlike auditory sensation, perception, working memory, and attention, ERPs that index language function have not yet been leveraged to study LiD. This is surprising given that these well-established procedures have been adopted for studying many other clinical populations (D’Arcy et al., 2011; Klumpp et al., 2010; Marchand et al., 2002; Olichney et al., 2008), and that difficulties in speech processing represent the primary complaint for this clinical population. The N400 ERP is linked to the processing of meaning and can be evoked using linguistic or nonlinguistic stimuli (e.g., pictures). Regardless, the amplitude of the N400 is strongly affected by stimulus predictability. The popular semantic anomaly paradigm involves analyzing the N400 with respect to the predictability of a word in a sensible sentence, which provides the necessary context for predicting future semantic input. It is larger for words that are less predictable (Kutas & Federmeier, 2011). The difference in amplitude between the N400s evoked by expected versus unexpected words is known as the N400 effect. The N400 is not a unitary neural entity. Rather, it is a label for stimulus-related brain activity with a sensitivity to linguistic meaning (Lau et al., 2008). Like recent behavioral tests aimed at measuring the contribution of language skills to LiD (Dillon et al., 2025; Zhou et al., 2026), the semantic anomaly paradigm is based on the Cloze procedure, which quantifies the predictability of words by measuring how accurately people are able to generate that word when it is omitted from a sentence (Taylor, 1953). While the N400 has not been studied yet in LiD, it has been used to investigate other pediatric populations with language or hearing impairments (Kallioinen et al., 2016; Pijnacker et al., 2017). It has also been examined as a potential method for passive speech-in-noise audiometry (Jamison et al., 2016).

Altogether, the behavioral and electrophysiological evidence in children with LiD support the need for comprehensive assessment, making the need for expedient procedures like Cheech apparent. Cheech can be implemented with any speech stimulus, but the amount of effort that people are willing to expend towards listening is thought to be gated by the value of the incoming signal to the listener; in other words, how motivated they are (Pichora-Fuller et al., 2016). Furthermore, differences in the cognitive demands imposed by isolated sentences versus continuous narratives have led some researchers to advocate for tests of speech comprehension that use longer, more naturalistic stimuli (Best et al., 2016). Thus, the implementation of Cheech for the present study used an engaging short story. By applying the Cheech method to a naturalistic narrative with designated target words, it is expected that the full range of auditory and cognitive ERPs described above should be observed. Based on recent literature that has strongly implicated cognition in LiD (Kojima et al., 2024; Magimairaj et al., 2020; McGrath et al., 2023; Petley et al., 2021, 2024; M. Sharma et al., 2014), it is further expected that cognitive ERPs like P300 and N400 will show robust differences between these groups.

## 2. Method

### 2.1 Participants

Thirteen children with LiD (8.1 – 14.7 years of age) and fifteen TD children (8.6 – 16.8 years of age) participated in this study, which took place at a single time point of an overarching longitudinal research program examining the pathology of LiD. All of the procedures regarding eligibility, recruitment, and behavioral testing were the same as for other reports from this research program (Hunter et al., 2020, 2023; Kojima et al., 2024; D. R. Moore et al., 2020; Petley et al., 2021, 2024; Stewart et al., 2022). In brief, eligibility criteria included the absence of any condition that would hinder test completion (neurological, psychiatric, or intellectual), English as a native language, and normal air conduction pure tone thresholds at the conventional frequencies (0.25 – 8 kHz; ≤ 20 dB HL). Children in the TD group were also excluded for reported developmental delays or learning or attention disorders. All study inclusion criteria, sociodemographic and health information were evaluated based on a structured background questionnaire completed by caregivers. Group inclusion was based on a reliable and validated caregiver questionnaire that is used to rate the presence of common symptoms of LiD (ECLiPS; (Barry & Moore, 2021; Denys et al., 2024) The study was approved by the Institutional Review Board of Cincinnati Children’s Hospital Research Foundation and all participants received monetary honorariums.

Demographics for the participants used in the present analysis, with respect to age, sex, and caregiver-reported listening difficulties near the time of data collection are summarized in Table 1.

**Table 1:** Demographics for the full study sample. Means and standard deviations are provided for age and ECLiPS Total Scaled Scores. All other values are frequencies.

|  | **TD** | **LiD** |
| --- | --- | --- |
| ***Number of Participants*** | 15 | 13 |
| ***Age*** | 11.8 (2.5) | 11.5 (1.8) |
| ***Sex*** |  |  |
| *Male* | 5 | 11 |
| *Female* | 10 | 2 |
| ***ECLiPS Total Scaled Score*** | 11.2 (2.7) | 2.9 (2.4) |

### 2.2 Procedures

Other procedures included in this overarching research design included neuroimaging using MRI, other EEG protocols, and a range of behavioral tests of auditory and cognitive function. Measures that are relevant to the present analysis are described in detail below.

#### 2.2.1 Caregiver-Reported Listening Difficulties (ECLiPS)

The ECLiPS questionnaire quantified all participants’ listening and communication abilities (Barry et al., 2015; Barry & Moore, 2021). The ECLiPS is a 38-item instrument that describes behaviors related to listening and communication in children along five subscales, three composite scores, and a total score. British data were used to provide age-scaled scores standardized for a population mean of 10 (SD = 3). Previous work has shown close congruence with U.S. (Petley et al., 2021) and Belgian (Denys et al., 2024) samples. Caregiver assessments on each item are provided on a five-point Likert scale (from “strongly agree” to “strongly disagree”). Participants in the LiD group had ECLiPS total scaled scores of less than 7 or an existing diagnosis of APD.

#### 2.2.2 Speech Listening in Noise (LiSN-S)

The Listening in Spatialized Noise – Sentences test (LiSN-S; Brown et al., 2010; Cameron & Dillon, 2007; Phonak/NAL, 2011) is used to evaluate the comprehension of a target speech stream in the presence of two competing talkers. The present study used the U.S. edition of the task, which was performed using a commercial CD (Phonak/NAL, 2011) via a laptop computer, a task-specific soundcard, and Sennheiser HD 215 headphones. During the LiSN-S, the participant repeats target sentences after they have been heard concurrently with the competing talkers. The benefit that the listener experiences from the provision of spatial separation and/or vocal differences between the target talker and the distractors is measured by completing the task under four different conditions: co-located identical voices, spatially separated identical voices, co-located different voices, or spatially separated different voices.

The target voice always remains at 0° azimuth, while the distractor voices move to ±90° azimuth when separated. Since the task is administered via headphones, these locations are simulated using generic head-related transfer functions (Humanski & Butler, 1988). Three scores, referred to as the Talker Advantage, Spatial Advantage, and Total Advantage, are derived through a subtraction process. These scores reflect the improvement of the participant’s speech reception threshold when spatial and/or vocal cues are provided. This subtraction process is thought to minimize the influence of cognitive factors like selective attention (Moore & Dillon, 2018). All analyses presented here are based on age-normed scores (Z-scores). Our previous work (Kojima et al., 2024; Petley et al., 2021) identified the Spatial Advantage and Talker Advantage scores as significant predictors of caregiver-reported listening skills on the ECLiPS.

#### 2.2.3 Cognition (NIH Toolbox Cognition Battery)

Participants’ cognitive abilities were evaluated using the NIH Toolbox Cognition Battery (Weintraub et al., 2013), which was administered online or via an iPad app, as indicated by Toolbox recommendations at the time of data collection, in a quiet room or sound-attenuated booth. The entire Battery involves up to eight standardized tests that measure specific aspects of fluid or crystallized cognition. Our previous work identified the Picture Vocabulary test (PVT), Dimensional Change Card Sort test (DCCS), and the Total Cognition score as significant predictors of listening skills (Kojima et al., 2024; Petley et al., 2021). The PVT adaptively evaluates a participant’s receptive vocabulary by presenting an audio recording of a word along with four pictures. The participant’s task is to select the picture that best matches the word. The DCCS measures one aspect of executive function: cognitive flexibility. Participants are presented with two cards that match a sample image along one of two possible dimensions, either shape or color. Unlike the classic Wisconsin Card Sorting Task (Berg, 1948), the dimension on which the cards should be sorted is explicitly specified, eliminating the need for inference, but it can change between trials, rendering the task sensitive to attention switching. The Total Cognition score is a composite derived from several tests, including five assessments of fluid cognition and two assessments of crystallized cognition. The fluid cognition tests include the DCCS, Flanker Inhibitory Control and Attention test (executive function and attention), Picture Sequence Memory test (a test of episodic memory), List Sorting Working Memory test, and Pattern Comparison Processing Speed test.

Tests of crystallized cognition include the PVT and the Oral Reading Recognition test (expressive language). The present analyses use age-normalized scores.

#### 2.2.4 Cheech Listening Task

The speech stimulus for the present study was a 16.9 minute story called “The Box of Robbers,” which is a chapter from the compilation “American Fairy Tales” by L. Frank Baum (Baum, 1901). An English narration of this story by a male speaker with a North American accent was obtained from the LibriVox public-domain audiobook repository (https://librivox.org). This story was converted to Cheech via frequency multiplexing. Specifically, the speech and energy-matched chirps were bandpass filtered to occupy non-overlapping portions of the frequency spectrum. The resulting Cheech story contained 17,058 chirps and comprised speech energy in the 0 – 250, 500 – 1,000, and 2,000 – 4,000 Hz frequency bands, and chirp energy in the 250 – 500, 1,000 – 2,000, and 4,000 – 10,000 Hz bands, see Figure 1. The filtered chirps were temporally aligned to glottal pulses in the speech to facilitate perceptual binding. A subset of these chirps were inserted with a longer post-chirp pause to permit the measurement of the MLR and LLAER. The resulting speech stream was somewhat robotic / noisy, but highly intelligible. Participants sat in a double-walled soundproof booth and listened to the story, which was delivered using Presentation® software (Neurobehavioral Systems, Inc., Berkeley, CA) diotically via Etymotic ER-3 insert earphones at 70 dB SPL. The word “and” was designated as the target for the word detection task, which participants identified by pressing the spacebar on a standard keyboard.

**Figure 1:**
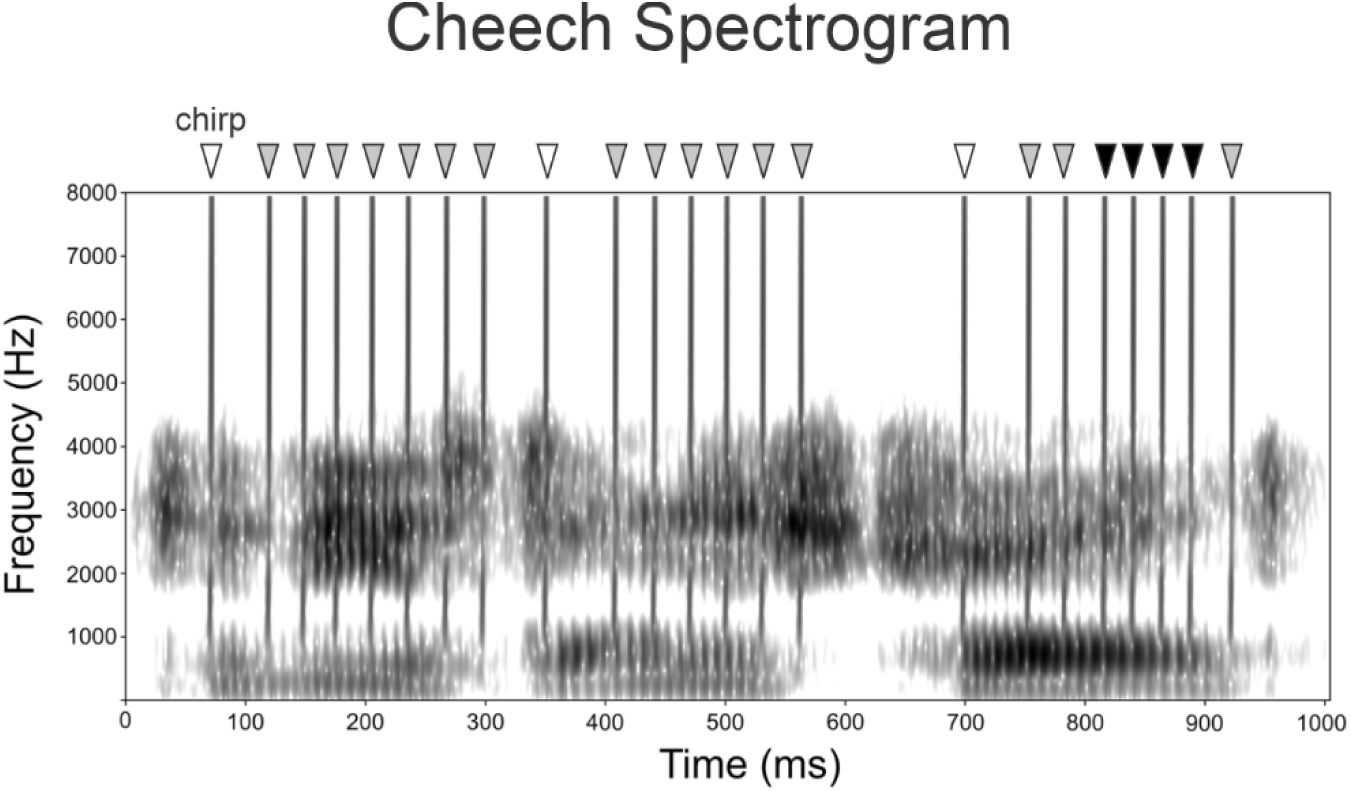
Spectrogram for a 1000 ms sample of Cheech. Note that chirp and speech energy occupy interleaved but separate bands of the frequency spectrum. Inverted triangles illustrate which chirp onsets might be used to derive the LLAER (white-filled), MLR (white-and grey-filled) and ABR (white, grey, and black), with later responses requiring longer pauses between chirps to permit their evolution.

#### 2.2.5 Electroencephalographic Recording

While participants listened to the story, EEG data were collected continuously using a 64-channel actiCHamp system (Brain Products, GmbH, Inc., Munich, Germany). Fifty-three electrodes were mounted in an elasticized cap with an equidistant layout arranged around a vertex sensor located at Cz of the 10-20 system. Additionally, single electrodes were placed below the right eye, on each mastoid, and on the tip of the nose, which served as the online reference. Impedances at all electrodes were maintained below 30 kΩ. EEG data were collected at 10 kHz and stored for offline analysis. In addition to the digital triggers that marked the onset of each chirp, an audio channel was also delivered to a StimTrack unit (Brain Products GmbH, Inc., Munich, Germany) connected to the auxiliary input of the EEG amplifier. Data recorded on this auxiliary channel were used to realign the digital triggers to the true chirp onsets.

#### 2.2.6 Supplementary Word Predictability Study

As previously described, the classic method for obtaining a word’s predictability in its context is the Cloze probability approach, which omits a single word from a sentence and quantifies the proportion of people who fill it in with a given word, as determined via survey (Taylor, 1953). Contrasting the neural response to words that match and mismatch the expected terminations of high Cloze probability sentences is a classic approach for deriving the N400 effect (Kutas & Hillyard, 1984). To emulate this procedure with the story used for the present analysis, two survey studies were conducted, in which the sentences of the story were presented one at a time, with the terminal words omitted. Each survey covered half of the “Box of Robbers,” with the full text for the first half of the story provided at the beginning of the second survey. Participants filled in the single word that they believed would best complete each sentence and were then presented with the correct terminal word. In this way, they could incorporate their knowledge of the story, as originally written, into their later predictions. Thirty-eight young adults, age 18 – 23 years (*M* = 19.2, 18 female) completed the first survey, and 34 young adults, age 16 – 23 years (*M* = 19.2, 15 female) completed the second. Participants were allowed to complete both surveys if desired. One participant in each study reported that English was not their native language. These studies were approved by the Clarkson University institutional review board and participants received course credit for their involvement. The survey data were used to identify high and low predictability sentence-terminal words from the story. A given word, or its synonyms, were produced by ≥ 67% or < 15% of respondents to qualify as a high or low predictability word, respectively.

### 2.3 Data Analysis

#### 2.3.1 Word Detection Task Performance

Responses to the target word “and” in the Cheech story were deemed correct if they occurred between 100 and 2000 ms from word onset.

#### 2.3.2 Evoked Responses

The EEG data were analyzed using a combination of custom scripts, EEGLAB v13.6.5b (Delorme & Makeig, 2004), and ERPLAB v8.0 (Lopez-Calderon & Luck, 2014), implemented in Matlab R2018b (Mathworks, Inc.). The data were first downsampled (following an anti-aliasing filter) as needed to derive each type of ERP. Downsampling to 2,000 Hz was required for the MLR, while 1,000 Hz was used for the LLAER, P300, and N400. The ABR did not require downsampling. The 1,000 Hz downsampled data were then visually inspected for channels with poor data quality. These channels and portions of the data that violated the assumption of stationarity for decomposition via independent component analysis (ICA), either with respect to amplitude (> 100 µV) or frequency (for example, strong, sporadic muscle artifacts) were rejected before performing ICA decomposition. Independent components were computed using Infomax ICA (implemented in EEGLAB) on average-referenced data that were band-pass filtered between 2 and 30 Hz using a 2^nd^ order Butterworth filter applied in the forward and backward directions (Klug & Gramann, 2021; Winkler et al., 2015). The resulting ICA matrices were then used to correct ocular and cardiac artifacts in the data for the LLAER, P300, and N400. Missing channels were interpolated when possible.

Filtering using 8^th^ order Butterworth filters applied in the forward and backward directions was applied using different cutoffs for several types of ERPs. The ABR required a passband from 100 to 1,500 Hz, the MLR required a passband from 15 to 200 Hz, and the LLAER required a passband from 0.5 to 20 Hz. Owing to their larger size and therefore lower frequency content, filtering for the P300 and N400 used a 2^nd^ order filter with a 0.5 – 20 Hz passband. The ABR and MLR were derived solely from the vertex (Cz), re-referenced to linked mastoids. The LLAER, P300, and N400 effect used all channels, re-referenced to an average reference. The continuous EEG data were then segmented into epochs. For the ABR, epochs were from-2 to 24 ms from the onsets of all chirps. For the MLR, the epochs extended from-5 to 60 ms from the onsets of any chirps that did not have another chirp in the 50 ms prior to or the 48 ms following their onset. The LLAER was obtained using epochs that extended from-50 to 500 ms from the onset of chirps that had no other chirps in the 150 ms prior to or the 48 ms following their onsets.

Additionally, to prevent overlap with processes related to target detection, linguistic processing, or movement, chirps that fell within 1000 ms following a target word onset, or 1000 ms prior to a response, as well as chirps occurring at the onset of the words used for the N400 analysis, were not used to compute the LLAER. The P300 and N400 used epochs from 50 ms prior to word onset to 950 ms after onset. To observe the P300, epochs were obtained for non-target phrase-onset words and correctly detected target words (hits). For the N400 effect, epochs were obtained for sentence-terminal words with high and low predictability, as determined by the supplementary word predictability study.

Averages were computed following the rejection of epochs containing artifacts. These artifacts were detected automatically using a +/-35 µV voltage threshold for the ABR and a +/-75 µV threshold for the MLR. For the LLAER, P300, and N400, the voltage threshold was +/-75 µV for most participants but was relaxed to +/-95 µV for three participants (one TD, two LiD) who demonstrated high-amplitude alpha on posterior channels. Descriptive statistics regarding the number of accepted epochs for all averaged waveforms (ABR, MLR, LLAER, P300, and N400 effect) and the number of ICA components rejected during artifact correction are reported in Supplementary Table 1. Latencies and amplitudes for the ABR, MLR, and LLAER were measured at Cz. The Cheech ABR differs from the classical click-evoked ABR in that the peaks prior to Wave V are not consistently observed, and it shows pronounced troughs prior to and following Wave V. Thus, both these troughs and Wave V were measured, with the pre-Wave V trough lying between 1 and 5 ms, Wave V emerging between 4 and 8 ms, and the post-Wave V occurring between 6 and 10 ms from chirp onset. For the MLR, the latency and amplitude of Na was measured at the largest negative peak between 10 and 27 ms from chirp onset, and Pa was measured as the largest positive peak between Na and 53 ms from chirp onset. For the LLAER, P1 was measured as the largest positive peak between 25 and 90 ms. N1 was only visible as a small trough between P1 and P2, therefore it was not measured. A sustained negativity (SN) was measured as the most negative peak between 200 and 480 ms. P2 was identified as the largest positive peak falling at least 50 ms after the P1 peak and 50 ms before the SN peak.

The P300 and N400 effect often have a complex wave shape, and their peaks are identified in difference waves between two stimulus conditions. The P300 peak was identified between 250 and 700 ms from chirp onset in a difference wave computed by subtracting the ERP to non-target phrase onset words out of the response to correctly detected target words. Since it is typically maximal at parietal or centro-parietal sites, it was measured between 250 and 550 ms at a midline parietal electrode (Ch5). The N400 effect was measured in a difference wave in which the ERP to high predictability words was subtracted from that evoked by low predictability words. It is typically maximal at parietal sites, so its latency and amplitude were measured at a slightly more posterior, midline parietal site (Ch47). All responses that involved a single, defined peak were measured as instantaneous peak amplitudes. These included the peaks and troughs of the ABR, Na and Pa of the MLR, and P1 and P2 of the LLAER. Responses with complex wave shapes, including the SN of the LLAER, P300, and N400 effect were measured as mean amplitudes in a ± 25 ms window surrounding the peak.

### 2.4 Statistical Analyses

All statistical analyses were carried out in R (version 4.0.2, the Foundation for Statistical Computing). To determine the influence of maturation, all ERP amplitudes and latencies, as well as accuracy and mean RT on the word detection task, were examined for a correlation with age using Spearman correlations. A similar correlational analysis was performed to examine the relationships between these variables and measures of cognition and auditory perception that our previous work identified as predictors of caregiver-reported listening abilities on the ECLiPS. Due to significant correlations amongst several measures of the ABR (see Supplementary Figure 1), Bonferroni corrections were applied in a familywise manner and adjusted *p*-values are reported for those variables. The threshold for significance for all tests was *p* ≤.05.

For many ERP components, including the peaks of the ABR and LLAER, and the N400 effect, the basis of LiD was explored via between-group comparisons of peak amplitudes and latencies. Due to the strong relationships amongst ABR measurements, and to maximize opportunities for direct comparison with other studies, group comparisons on these measures were limited to the amplitude and latency of Wave V. These comparisons, as well as those for performance on the word detection task, were performed using Student’s *t*-tests when the assumptions of normality and homogeneity of variance were met, Welch’s *t*-tests when only normality was met, and Wilcoxon rank sum tests when normality was not met. Normality was tested via Shapiro-Wilk normality tests as implemented in base R. Homogeneity of variance was evaluated using the Levene’s Test function of the *car* package (Fox & Weisberg, 2019). The groups that were used for these comparisons were age-matched, but the TD group generally had a greater proportion of females and a higher degree of maternal education than the LiD group. Effect sizes are reported as Cohen’s *d*, computed using the *effsize* package (Torchiano, 2020).

The P300 and N400 effect reflect cognitive differences in processing between two different experimental conditions. In the case of the P300, the increased positive voltage obtained for hits versus non-target stimuli reflects the selective attention and working memory updating processes involved in target detection (Polich, 2007). Accuracies on the word detection task tended to be poor, so this response could only be measured in a small subset of participants. Its presence was verified via a repeated measures Student’s *t*-test comparing the mean amplitude at the time of the P300 peak between non-target words and hits, using participants from both groups. For the N400 effect, a greater negative voltage should be observed for unpredictable versus predictable words, reflecting a process of semantic integration (Kutas & Federmeier, 2011). Separate Student’s *t*-tests were performed for the LiD and TD groups comparing the mean amplitude at Ch47 between the low-and high-predictability word conditions. For these repeated measures, Cohen’s *d* was computed using the *rstatix* package (Kassambara, 2020).

Owing to the highly unusual scalp distribution observed for the N400 effect in the LiD group, group comparisons for N400 amplitude of the low minus high predictability waveform were performed at a pooled frontopolar site (Fp; Ch10 and Ch40).

## 3. Results

### 3.1 Relation to Cognition and Auditory Perception

There were several meaningful relationships between the ERP and behavioral measures obtained from this task and scores on standardized tests of auditory perception and cognition. Figure 2 illustrates the correlations that reached significance. Most of these were observed for the NIH Toolbox Cognition Battery. The DCCS task correlated significantly with the latency of the pre-Wave V trough of the ABR, *r_s_*(26) =-0.46, adj. *p* =.039 (adjusted for significant correlations with Wave V and post-Wave V latencies) and accuracy on the word detection task, *r_s_*(26) = 0.51, *p* =.006. Accuracy was also significantly related to Total Composite scores, *r_s_*(26) = 0.51, *p* =.005. Finally, Total Composite scores also exhibited a significant relationship with the amplitude of the N400 effect, *r_s_*(22) =-0.40, *p* =.050. Participants with better scores on the DCCS had earlier pre-Wave V troughs and superior accuracy on the word detection task. Those with higher Total Composite scores showed earlier pre-wave V troughs and larger N400 effects (note that the negative correlation results from the negative polarity of the N400 effect). Two ERP measures demonstrated significant relationships with the LiSN-S Spatial Advantage score: P1 latency, *r_s_*(26) =-0.39, *p* =.041 and P2 amplitude, *r_s_*(26) =-0.48, *p* =.010. Owing to a significant relationship with age (see Supplementary Figure 2 and Supplementary Table 2), which indicated that older participants had smaller and earlier P1 responses, only age-corrected P1 latencies and amplitudes were used to explore these correlations. Participants with better Spatial Advantage scores tended to have earlier P1 latencies and smaller P2 amplitudes.

**Figure 2:**
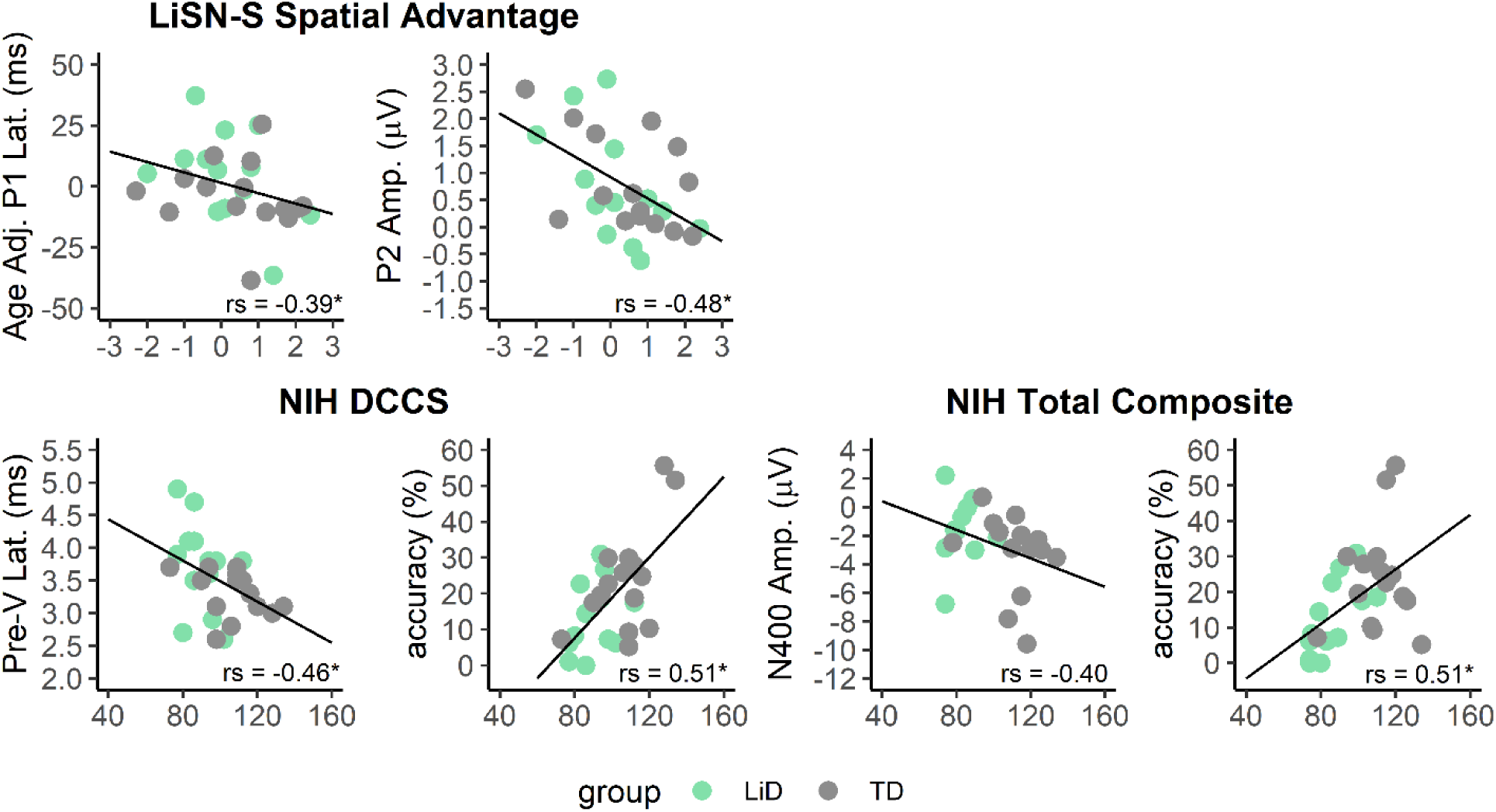
Significant correlations between ERP and behavioral measures from the Cheech listening task and measures of auditory perception and cognition. Accuracy refers to target word detection. * *p* <.05

### 3.2 Early Responses: ABR and MLR

Figure 3A illustrates the ABR at Cz / Ch1. A highly-synchronized Wave V, with adjacent troughs, is evident for both groups. Table 2 provides the descriptive statistics for all ERP measurements. Only the amplitude and latency of Wave V were compared between the groups. The response tended to be smaller for LiD than TD participants, *t*(24) = 1.73, *p* =.097, *d* = 0.68 and later as well, *t*(24) = 1.74, *p* =.094, *d* = 0.68, but neither effect reached significance. All group comparisons are summarized in Supplementary Table 3. The components of the MLR, shown in Figure 4B, could not be compared between groups due to the large proportion of participants who demonstrated a posterior auricular muscle (PAM) reflex artifact, particularly in the TD group. Examined across all participants who showed no PAM reflex artifact, regardless of group (5 TD and 9 LiD participants), Table 2 demonstrates that both Na and Pa were robust, since the 95% confidence intervals for their amplitudes did not cross zero.

**Figure 3:**
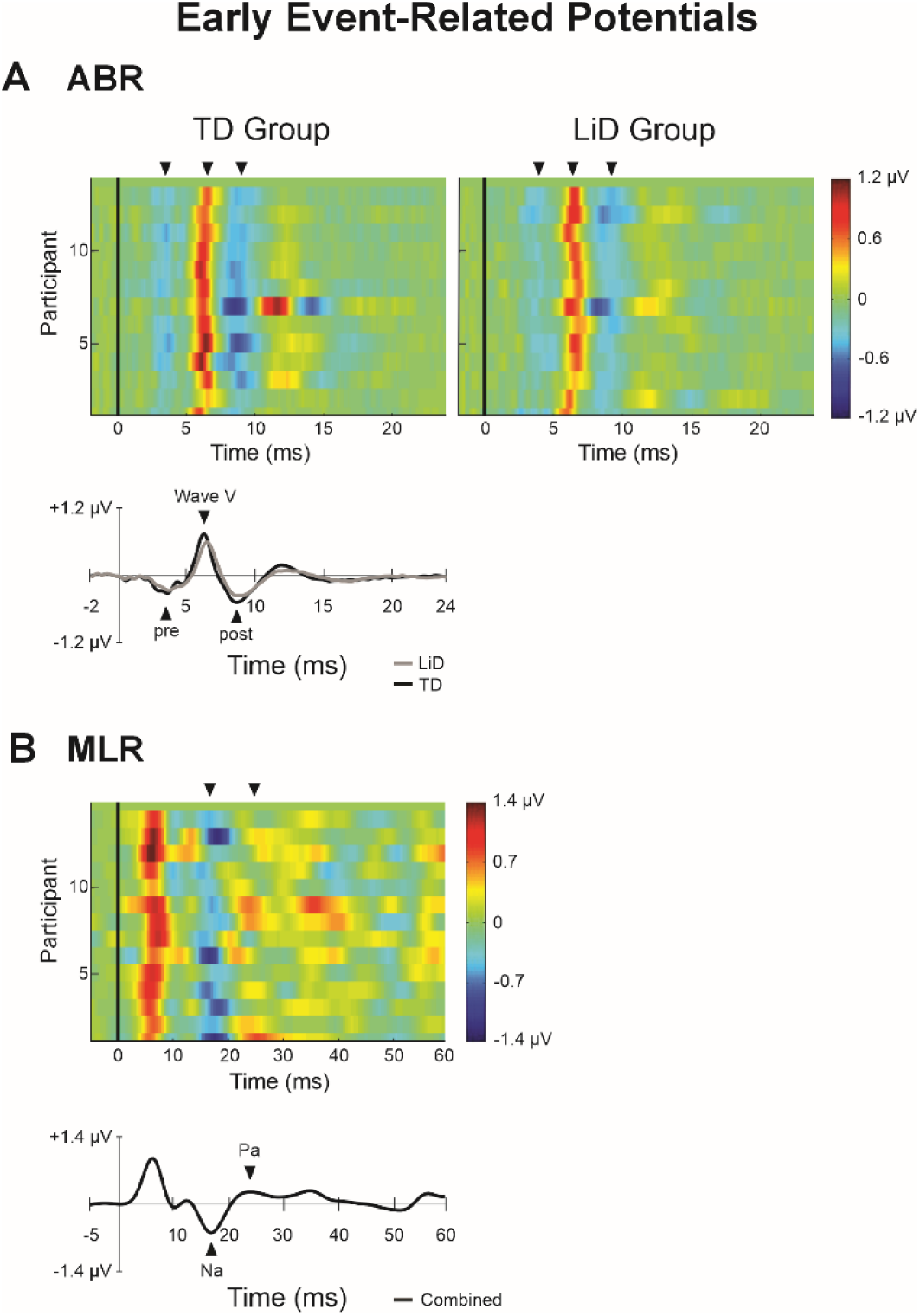
Early ERPs including the ABR and MLR. ERP images reveal similarities in timing for the response across participants, with different participants in each row and voltage indicated via heatmap. Grand averages are found below the ERP images. Unlike the ABR, the MLR could not be derived separately for the groups due to the prevalence of PAM reflex artifacts.

**Figure 4:**
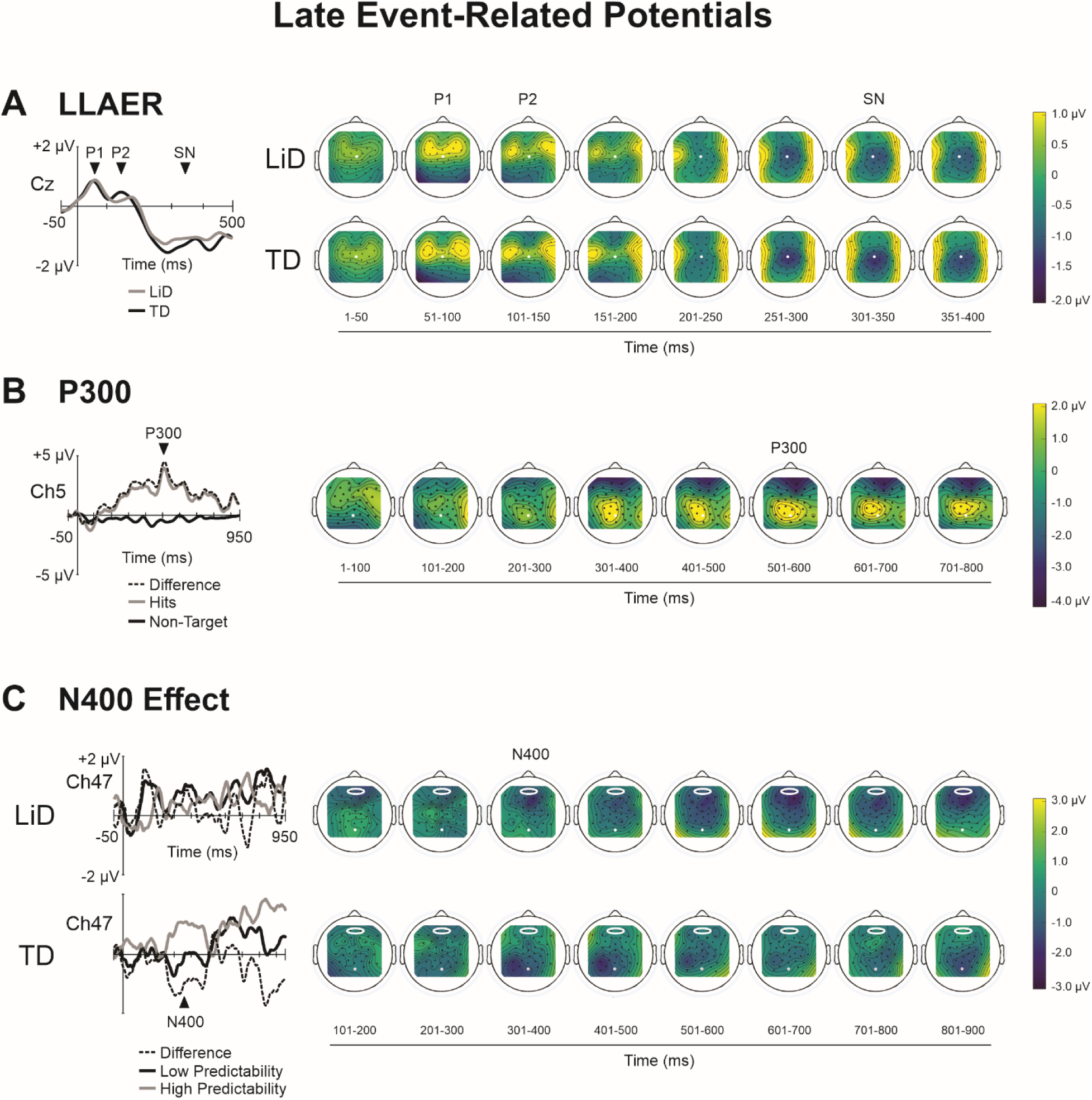
Grand averages and scalp distributions for the long-latency ERPs. White electrode positions and encircled electrode pools indicate the sites of measurement. All other electrode positions are shown in black. For the LLAER and N400 effect, TD participants are shown in the top row while those in the LiD group are on the bottom. Due to low rates of target word detection, the P300 includes participants from both groups.

**Table 2:** Descriptive statistics for all ERP measurements that could be obtained separately for the TD and LiD groups. P1 amplitude and latency are shown as their original values, without age adjustment.

|  | TD |  |  | LiD |  |  |
| --- | --- | --- | --- | --- | --- | --- |
|  | N | Mean | 95% CI | N | Mean | 95% CI |
| **ABR (Cz/Ch1)** |  |  |  |  |  |  |
| Pre-Wave V latency (ms) | 13 | 3.32 | 3.12, 3.51 | 13 | 3.72 | 3.35, 4.10 |
| Pre- Wave V amplitude ($\mu V$) | 13 | -0.36 | -0.41, -0.31 | 13 | -0.31 | -0.35, -0.27 |
| Wave V latency (ms) | 13 | 6.28 | 6.17, 6.40 | 13 | 6.45 | 6.30, 6.61 |
| Wave V amplitude ($\mu V$) | 13 | 0.82 | 0.71, 0.93 | 13 | 0.70 | 0.62, 0.78 |
| Post-Wave V latency (ms) | 13 | 8.59 | 8.47, 8.72 | 13 | 8.80 | 8.59, 9.01 |
| Post-Wave V amplitude ($\mu V$) | 13 | -0.51 | -0.61, -0.42 | 13 | -0.41 | -0.48, -0.35 |
| **LLAER (Cz/Ch1)** |  |  |  |  |  |  |
| P1 latency (ms) | 13 | 53.2 | 44.5, 61.8 | 13 | 63.5 | 53.3 |
| P1 amplitude ($\mu V$) | 13 | 1.05 | 0.58, 1.51 | 13 | 1.06 | 0.68, 1.44 |
| P2 latency (ms) | 13 | 151.7 | 137.0, 166.4 | 13 | 144.5 | 124.5, 164.5 |
| P2 amplitude ($\mu V$) | 13 | 0.79 | 0.31, 1.26 | 13 | 0.74 | 0.17, 0.31 |
| SN latency (ms) | 13 | 339.9 | 297.7, 382.2 | 13 | 350.6 | 305.5, 395.7 |
| SN amplitude ($\mu V$) | 13 | -1.69 | -2.23, -1.14 | 13 | -1.48 | -1.92, -1.05 |
| **N400 Effect** |  |  |  |  |  |  |
| Latency (ms) | 10 | 369.0 | 321.9, 416.1 | 10 | 389.7 | 318.8, 460.6 |
| Amplitude at Ch47 ($\mu V$) | | | | | | |
| Low Predictability | 10 | -1.25 | -2.36, -0.14 | 10 | -0.65 | -1.35, 0.06 |
| High Predictability | 10 | 1.32 | 0.71, 1.93 | 10 | 0.96 | -0.57, 2.50 |
| Amplitude at Fp ($\mu V$) | | | | | | |
| Difference (Low – High) | 10 | 0.66 | -0.27, 1.60 | 10 | -0.81 | -1.85, 0.22 |

### 3.3 LLAER

The LLAER is illustrated in Figure 4A. In light of the significant age correlations observed for the latency and amplitude of P1, these measures were age-corrected prior to group comparisons. No measures of the LLAER differed significantly between the two groups.

### 3.4 Target Word Detection and P300

Regardless of group membership, participants tended not to detect the target word (TD group accuracy *M* = 21.3%, *SD* = 13.3%; LiD group *M* = 12.3%, *SD* = 10.3%), but there was a trend towards better accuracy for the TD group, *t*(24) = 1.92, *p* =.066, *d* = 0.77. Group differences in RT did not approach significance (see Supplementary Table 3). As a result of this poor rate of target detection, few participants had enough hits to permit measurement of the P300. The response, shown in Figure 4B, was therefore measured by combining participants across the groups. Despite this challenge, the comparison between the non-target and hit waveforms indicated that P300 response was robust at Ch5, *t*(5) = 4.22, *p* =.008, *d* = 1.7, with a more positive response to hits than non-target words. Like the MLR, this effect is summarized in Table 3.

**Table 3:** Descriptive statistics for all ERP measurements that were obtained across the groups.

|  | N | Mean | 95% CI |
| --- | --- | --- | --- |
| **MLR (Cz/Ch1)** |  |  |  |
| *Na latency (ms)* | 14 | 17.0 | 16.5, 17.6 |
| *Na amplitude ($\mu V$)* | 14 | -0.69 | -0.85, -0.53 |
| *Pa latency (ms)* | 14 | 29.4 | 25.9, 32.8 |
| *Pa amplitude ($\mu V$)* | 14 | 0.48 | 0.38, 0.58 |
| **P300 (Ch5)** |  |  |  |
| *Difference wave latency (ms)* | 6 | 520.8 | 454.0, 587.6 |
| *Non-Target amplitude ($\mu V$)* | 6 | -0.18 | -0.71, 0.35 |
| *Missed Target amplitude ($\mu V$)* | 6 | 0.56 | -0.79, 1.91 |
| *Detected Target amplitude ($\mu V$)* | 6 | 4.21 | 2.56, 5.87 |

### 3.5 N400 Effect

The N400 effect, in which the response to less predictable words is negative compared to predictable words, is illustrated in Figure 4C, and its amplitude is summarized in Table 2. The scalp distributions derived from the low minus high predictability difference wave clearly demonstrate a topographical difference for the response between TD and LiD participants. While the TD group showed a conventional, parietal maximum, the LiD group demonstrated a distinctly frontal distribution.

Accordingly, the difference in amplitude between the low-and high-predictability word conditions at Ch47 was significant for the TD group, *t*(9) = 3.59, *p* =.006, *d* = 1.13, but not the LiD group, *t*(9) = 2.09, *p* =.066, *d* = 0.66. A comparison between the two groups for the amplitude of the N400 effect in the low minus high predictability difference wave at the pooled frontopolar site very closely approached significance, with a large effect size for the difference between the groups, *t*(18) = 2.08, *p* =.052, *d* = 0.93.

## 4. Discussion

Like many other disorders of the central nervous system, the exact pathophysiology of APD and LiD remains unknown. When their symptoms are reported, clinicians are faced with a myriad of assessment options and little guidance as to which are indicated, particularly with respect to electrophysiology. The present results demonstrate that enriching natural speech with appropriately timed chirps, in a method called “Cheech,” permits concurrent measurement of many of the recommended ERPs for APD, spanning the speech processing hierarchy. They also provide indications as to which responses might be most valuable for differentiating children with LiD from their TD peers.

To avoid conflating pathophysiology with maturational effects, an important first step in evaluating group differences in these evoked responses was to establish possible correlations with age. Only one component, P1, was related to age, becoming earlier and smaller in older participants. This result is consistent with the literature on maturation of the LLAER using both speech (A. Sharma et al., 1997) and non-speech stimuli (Ponton et al., 2002). All other responses evoked by Cheech appear to be stable within the age range examined here (8-17 years of age).

Compromised auditory perceptual and cognitive abilities are well-established for children with LiD (Kojima et al., 2024; McGrath et al., 2023; Pascoinelli et al., 2021; Petley et al., 2021, 2024; M. Sharma et al., 2014), and correlational analyses revealed that these characteristics are related to the ERP measures yielded by Cheech. With respect to auditory perception, the LiSN-S Spatial Advantage score, which reflects the ability to use spatial separation to listen to a target voice in the presence of competing speech, was significantly correlated with both the age-adjusted latency of P1 and the amplitude of P2. Its correlation with the age-adjusted amplitude of P1 also approached significance (*p* =.063; see Supplementary Table 2). P1 and P2 are the dominant features of the neural response to discrete speech stimuli, like syllables (Čeponienė et al., 2005). Studies of envelope tracking using continuous speech have also yielded peaks with similar timing (Di Liberto et al., 2015; Zion Golumbic et al., 2013). For adults, effortful auditory attention (known as “listening effort”) is associated with greater synchronization of responses in this latency range, leading to clearer N1 and P2 components (Strauss et al., 2010). Thus, the weaker responses observed here for participants who score higher in spatial listening may reflect that they find listening to speech generally less effortful.

The correlational analysis also revealed many relationships between Cheech ERPs and cognition.

Perhaps surprisingly, a brainstem-level response, the latency of the pre-Wave V trough, showed significant correlations with scores on the NIH Dimensional Change Card Sort test. The pathways that generate the ABR involve few synapses, and the interpeak interval between waves III and V have been used to study changes in axonal conduction speed during maturation (Ponton et al., 1996). Considerable evidence also suggests an important relationship between CNS myelination and cognitive function (Fields, 2008). Thus, this feature of the ABR might correlate with cognition because it serves as a measure of myelination and axonal conduction speed. This study is not the first to identify a relationship between ABR measures and cognition. Both Wave V latency and amplitude have recently emerged as useful predictors of cognitive function in the elderly, independently of hearing level and age (Hamza et al., 2024). ABR-based measurements are also known to differentiate TD children from those with autism and learning disabilities (Pillion et al., 2018; Purdy et al., 2002). Very recent work using a Cheech approach similar to the present investigation has also shown better speech attention and comprehension for participants who show earlier and larger Wave V peaks (Mankel et al., 2024).

Unlike the ABR, the N400 effect directly reflects cognitive, though implicit, operations. Its amplitude correlated significantly with the NIH Total Composite score and very closely approached significance when examined for a relationship with vocabulary size (NIH PVT; *p* =.057, see Supplementary Table 2). As such, its relationship with behavioral measures of cognition was somewhat nonspecific. Further, all correlations were negative, indicating that better cognitive performance was associated with a more negative (i.e., larger) N400 effect. Of course, the fact that measures of cognition tend to correlate with one another is among the most well-established of all psychometric phenomena (Spearman, 1904).

When evoked in semantic anomaly paradigms, or using longer narrative stimuli as in the present study, the N400 effect reflects predictive mechanisms that pre-activate the memory representations of likely future input based on contextual information arising at multiple levels, from simple word meanings (lexico-semantics) to knowledge of the world and pragmatic reasoning (Kutas & Federmeier, 2011).

Since all levels of information may not yield the same predictions, the underlying mechanism of the N400 must flexibly rely on the information has the greatest predictive value at a given time (Dave et al., 2021). Beyond semantic knowledge as a component of cognition, the present results may therefore suggest a relationship between this implicit capacity for flexible prediction and cognition, broadly defined.

Our results also point to which ERP components might be most valuable in identifying children with LiD using the Cheech method. Overall, the response that most clearly differentiated these two groups was the N400 effect. Given the growing evidence for cognitive deficits as a component of LiD (Kojima et al., 2024; Magimairaj et al., 2020; McGrath et al., 2023; Petley et al., 2021, 2024; M. Sharma et al., 2014) this finding aligns with the observed correlation between the N400 effect and general cognition. Longer materials, like the story used here, provide information across multiple temporal scales, and accurate predictions may at times rely on facts that were learned several minutes earlier. The observation of a robust N400 effect in TD children suggests that this process is relatively effortless for them: accurate predictions of sentence-terminal words were generated, and violations of these expectations led to larger N400 responses. By contrast the N400 effect only approached significance for children with LiD. This lack of effective pre-activation may reflect an overall deficit in the predictive mechanisms that facilitate speech perception.

Children with LiD also demonstrated an unusual, frontal topography for the N400 effect. A between-group comparison for the size of this response in the frontopolar region very nearly reached significance and demonstrated a large effect size. While the topography of the N400 effect can vary depending on the stimulation modality (Kutas & Federmeier, 2011), a frontal distribution is atypical for this response. Instead, this distribution is characteristic of a similar response known as the FN400. The functional differences between the FN400 and N400 are a matter of debate (Bridger et al., 2012; Voss & Federmeier, 2011), with some proposing that the FN400 reflects a recognition process based on familiarity, thus explaining why it is typically observed during tasks that involve distinguishing old from new items (Bader et al., 2023). The crucial difference between the circumstances that evoke the N400 versus FN400 is whether the participant has intentionally adopted a strategy of episodic retrieval.

Following this rationale, it is possible that listeners with LiD compensate for their difficulties by engaging in an active comparison of incoming speech to what they heard previously.

Ours is the first study to examine the N400 effect in people with LiD, but Stewart and colleagues (2022) used functional MRI to examine the brain’s language networks using a sample of participants that overlapped with the current analysis. Their results revealed altered resting-state functional connectivity between brain regions that were activated when listening to speech for children with LiD. These differences only arose for regions involved in speech intelligibility and semantics. Group differences did not emerge for regions involved in basic aspects of speech processing like phonology. The present results are consistent with these differences across the speech processing hierarchy. Earlier ERPs that are associated with auditory perception, like P1 and P2 demonstrated no differences between the two groups, while the N400 effect did.

Behavioral research also points to altered language skills as potential contributors to LiD. It is well established that speech in noise thresholds for sentences are improved by valid semantic and grammatical information, which can be used to fill in segments of the speech stream that were missed (G. A. Miller & Isard, 1963). In a similar vein, listeners with larger vocabularies and faster lexical access generally possess superior speech in noise thresholds (Carroll et al., 2016; Kaandorp et al., 2016), and an increased vocabulary size supports speech recognition in noise for hard of hearing children with hearing aids relative to non-users of hearing aids (Walker et al., 2019). A previous analysis by our group using a sample that overlaps with the present study also identified NIH PVT scores as an important predictor of caregiver-reported listening skills (Petley et al., 2021). Although APD has been considered to be clinically distinct from developmental language disorders (DLD), LiD are often observed for children with DLD. These clinical populations are indistinguishable from one another on caregiver reports of listening skills as well as many assessments of auditory processing and speech intelligibility (Ferguson et al., 2011). Owing in part to the increased recognition of possible linguistic contributions to LiD, a clinical test has recently been devised to measure semantic inference in a similar manner to the paradigm used here to measure the N400 (Zhou et al., 2026). Given the sensitivity of many language tests to nonlinguistic cognitive factors like working memory and executive function, some researchers suggest that tests of receptive language may better reflect the demands that are imposed by everyday listening than those for expressive language (Falcone et al., 2026). By eliminating the need for an explicit response, an N400-based assessment has the potential to reduce the influence of nonlinguistic cognitive factors even further.

No other ERP statistically differentiated children with LiD from their TD peers, but trends towards smaller and later Wave V peaks were observed for children with LiD. While brief, non-speech stimuli like clicks and tones are typical for evoking the ABR, it can also be obtained in response to speech, and there appears to be considerable independence between the two types of measurements (Song et al., 2006). The Cheech stimulus employed here is a compound speech and chirp stimulus. As such, it is difficult to know whether results obtained using speech or non-speech stimuli provide the best points for comparison.

With respect to the click-evoked ABR literature, the present results mirror some of those obtained by Gopal (2002), who found that Wave V was smaller and later in children with auditory processing difficulties compared with their TD peers. Jirsa (2001) also observed prolonged Wave Vs in children with APD using maximum length sequence (extremely rapid) stimuli, but not all studies agree. Ankmnal-Veeranna and colleagues (2019) observed significantly prolonged click ABR Wave V peaks for children with suspected APD compared to adults, but not when compared to TD children. Allen and Allan (2014) obtained abnormal click ABR Wave V amplitudes (measured as a ratio vs. Wave I) and prolonged latencies in some children with suspected APD, but there were no significant group differences. By contrast, Hunter and colleagues (2023), using a sample that overlapped with ours, demonstrated earlier click-evoked ABR wave III and V latencies for children with LiD. Yet other studies have shown no differences in the click-evoked Wave V for children with LiD (Filippini & Schochat, 2009; Omidvar et al., 2023), but one of these also measured speech-evoked responses and found Wave V delays for their LiD group (Filippini & Schochat, 2009). Others have failed to confirm this finding, and instead show altered timing for other speech ABR peaks (Omidvar et al., 2023; Rocha-Muniz et al., 2012). Since difficulties with noisy or degraded speech are often considered to be the defining feature of LiD, speech-evoked ABRs may have special relevance for this population.

Regrettably, some of the ERPs that were evoked using Cheech could not be compared between the two groups. Na and Pa of the MLR could only be measured in a subset of participants due to frequent PAM reflex artifacts. Interestingly, these tended to affect a greater proportion of TD (10/15) than LiD (4/13) participants, though this difference did not reach significance, χ^2^(1, N = 28) = 2.30, *p* =.130. PAM reflex artifacts are a well-known hazard when measuring MLRs and are easily evoked by loud sounds (Bell et al., 2004). They are observed more readily to chirps than clicks, and their likelihood increases when the neck muscles are tense (Picton, 2010). The results of the present study suggest that every possible precaution should be taken to avoid PAM reflex artifacts in future studies using Cheech, including potentially decreasing the intensity of the stimulus, using a linked earlobe reference, and performing data collection with participants in a reclined position to reduce neck muscle tension.

Like the MLR, the P300 could not be compared between groups. The P300 occurs in response to actively-detected targets, and is thought to reflect the updating of stimulus representations in working memory following target detection (Polich, 2007). Since its amplitude is related to the amount of attention that is dedicated to incoming stimuli, alterations of this response are commonly used to make inferences regarding attentional function, for example in patients suffering from brain injuries (Petley et al., 2018; Sculthorpe-Petley et al., 2015) and mental illnesses like schizophrenia (Jeon & Polich, 2003). The experimental approaches that have been used to evoke the P300 for this purpose are too varied for succinct description, but oddball paradigms consisting of physically or temporally divergent targets presented unpredictably within trains of otherwise homogeneous stimuli are extremely common. Despite its popularity and the diversity of circumstances under which the P300 is studied, measuring it in response to target words in a continuous narrative is not an established procedure.

Perhaps the most similar example of P300 measurement with narrative stimuli is the work of Dwivedi and Gibson (2017), who used a noun (the word “tree”) as a target in visually-presented sentences (see also Selvanayagam et al., 2019). Their target word was marked by its font color, making its detection entirely independent of linguistic processing. Although the present approach was novel, for the subset of participants who detected the target word often enough to measure the response, the P300 had a typical centroparietal distribution and was robust. This suggests that the general procedure is sound, but that the target word requires optimization. Due to its frequent occurrence, the word “and” was selected as the target for the present study so that participants would not go long periods without hearing it.

Unfortunately, this property was insufficient to promote successful detection. One possible reason may be the semantic value of the word. “And” is a conjunction word, so it has little meaning on its own. Instead, it describes the relationship between other words. It may be that conjunction words capture relatively little attention when listening for the overall meaning of a narrative. Although performance was poor for all participants, there was a near-significant trend towards better accuracy for TD than LiD participants (*p* =.066). It may be that the extra effort that participants with LiD had to expend towards understanding the story limited their ability to perform the target detection task at the same time.

While the procedure for measuring the P300 in a narrative requires optimization, there are compelling reasons to do so. Smaller (Jirsa, 1992; Jirsa & Clontz, 1990; Mattsson et al., 2019) and/or delayed (Jirsa, 1992; Jirsa & Clontz, 1990; Krishnamurti, 2001) P300 responses have been observed for people with LiD on auditory target detection tasks using simple, nonverbal stimuli. Recent work with targets defined by changes in amplitude modulation rate, using a sample that overlaps with the present study, also showed reduced responses at parietal sites in the latency range associated with P300 when poorly-performing participants with LiD were included in the analysis (Petley et al., 2024). Finally, the response has been frequently been used to monitor changes in auditory processing following interventions for APD, largely revealing a decrease in latency over time and, to a lesser extent, increased amplitudes or emergence of the response when it was previously absent (Hajimohammadi et al., 2025). Insights from behavioral research also provide an impetus for P300 measurement, since attentional difficulties represent yet another possible factor contributing to LiD (Moore et al., 2010; M. Sharma et al., 2019). Since longer passages, like short stories, emulate the cognitive demands of real-world listening better than brief speech stimuli like words or sentences, future work should identify the best types of targets for evoking the P300 in such contexts.

## 5. Conclusions

Altered cognition is increasingly being recognized as a component of LiD, but central auditory processing remains an important aspect of assessments for patients who report such difficulties. By combining chirps with natural speech, Cheech makes it possible to objectively measure responses along the speech processing hierarchy, from central auditory processing through to cognitive operations like attention, working memory, and language, in a single session during which the participant can listen to an engaging story. Most of the ERPs measured by Cheech are unaffected by maturation in childhood and adolescence, and some show relationships with aspects of cognition, including vocabulary size, cognitive flexibility, and general cognition. More importantly, among these responses, the N400 effect clearly differentiated children with LiD from their TD peers, implicating language function as an important component of LiD. This finding dovetails with recently developed tests of linguistic prediction for use in their assessment. Further research to study LiD using optimized Cheech methods that eliminate PAM reflex artifacts and improve target word detection rates would improve the neural characterization of LiD, paving the way for diagnostic applications.

## Data Availability

All data produced in the present study are available upon reasonable request to the authors.

## 6. Acknowledgements

This research was supported by grant DC014078 from the National Institute of Deafness and other Communication Disorders (to DRM), by the Cincinnati Children’s Research Foundation, and by the office of the Assistant Secretary of Defense for Health Affairs through the Congressionally Directed Medical Research Program (CDMRP) Hearing Restoration Research Program (HRRP) under Award No. W81XWH-20-1-0485 (to LMM). DRM was supported in part by the NIHR Manchester Biomedical Research Centre. Opinions, interpretations, conclusions, and recommendations are those of the authors and are not necessarily endorsed by the Department of Defense. The authors also wish to acknowledge the assistance of Noah Campagna and Thu Nguyen in the collection of these data.

## 7. Data Availability

Data will be made available on reasonable request to the authors.

## Supplementary Figures and Tables

**Supplementary Figure 1:**
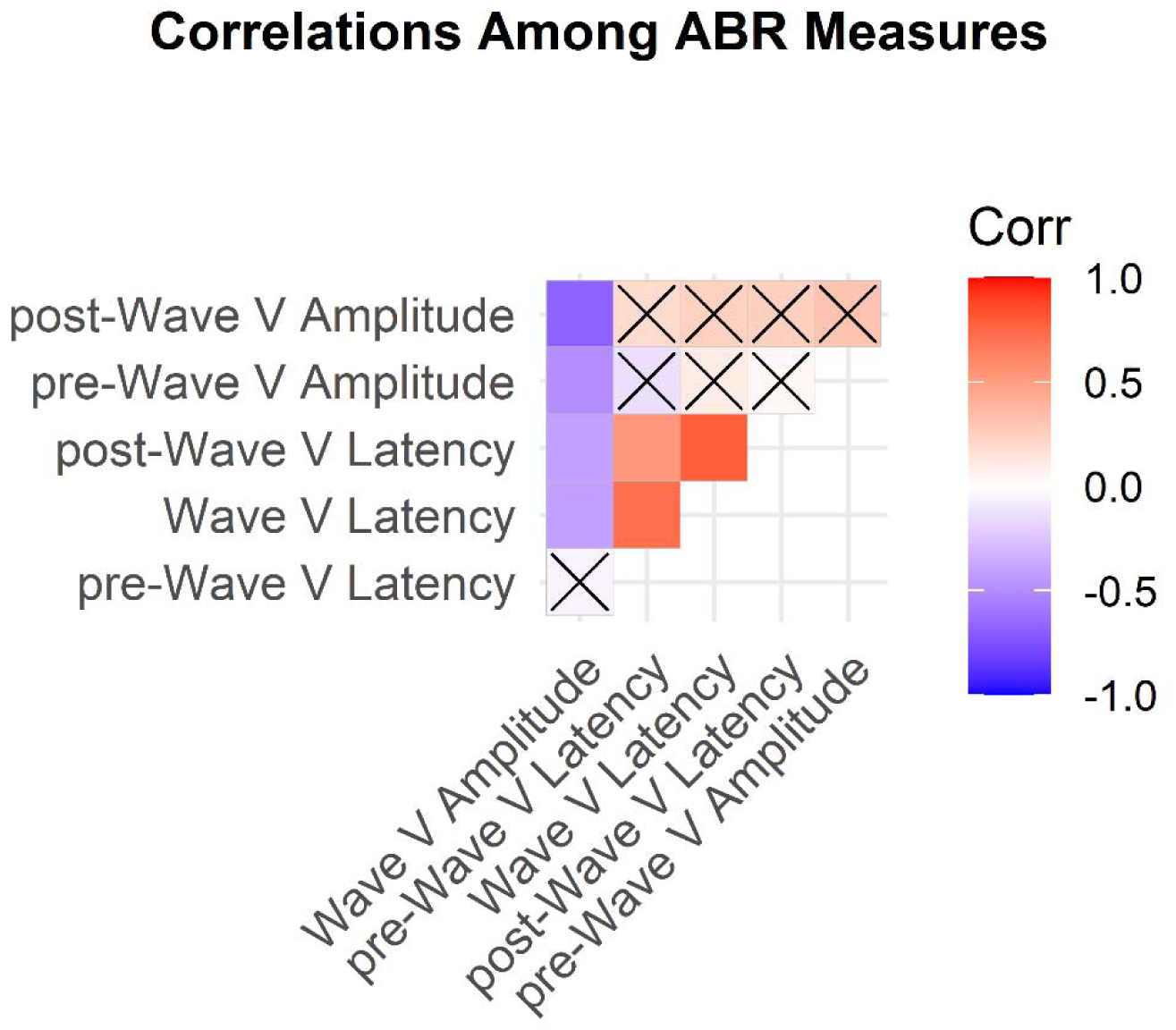
Correlations among all amplitude and latency measures for the ABR, including those for the pre-Wave V trough, Wave V, and the post-Wave V trough. Correlations that did not reach statistical significance (*p* >.05) are crossed out.

**Supplementary Figure 2:**
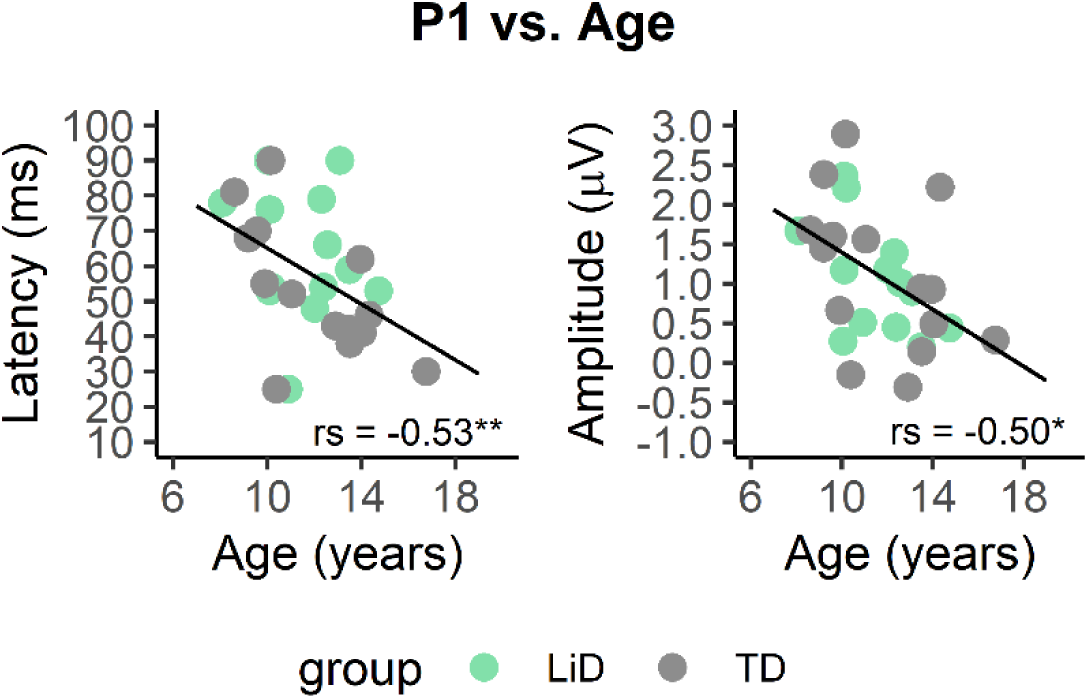
Significant correlations with age were observed for the latency and amplitude of P1: latency *r_s_*(26) =-0.53, *p* =.004, amplitude, *r_s_*(26) =-0.50, *p* =.007. No other ERP measures correlated age, nor did word detection accuracy or RT. Results for all age correlations that approached significance (*p* < 10) are reported in Supplementary Table 2.

---

**Supplementary Table 1:** EEG denoising and ERP data quality measures. All values are reported as mean *(SD)*. When data were combined across groups (as for the MLR and P300), values are only provided in the TD column.

|  | TD | LiD |
| --- | --- | --- |
| **ICA Components Rejected** | 3.5 (1.4) | 3.8 (1.3) |
| **Trials Accepted for Averaging** |  |  |
| ABR | 16979.9 (126.9) | 16716.2 (290.1) |
| MLR | 1358.1 (20.7) | - |
| LLAER | 485.8 (25.6) | 472.5 (27.7) |
| P300 (hits) | 36.2 (12.0) | - |
| P300 (misses) | 56.5 (10.4) | - |
| P300 (non-targets) | 512.5 (18.6) | - |
| N400 (low predictability) | 72.3 (3.1) | 68.3 (5.6) |
| N400 (high predictability) | 60.4 (3.1) | 57.6 (4.8) |

**Supplementary Table 2:** Summary of all correlational results that approached significance (*p* <.10). Adjusted *p*-values are reported for all ABR measures to account for correlations amongst them.

| Variable 1 | Variable 2 | *N* | *r<sub>s</sub>* | *p* |
| --- | --- | --- | --- | --- |
| Age | Na Amplitude | 14 | -0.47 | .087 |
|  | P1 Latency | 28 | -0.53 | .004 |
|  | P1 Amplitude | 28 | -0.50 | .007 |
|  | P2 Latency | 28 | 0.34 | .076 |
|  | P2 Amplitude | 28 | -0.35 | .071 |
| LiSN-S Spatial Advantage | Age Adj. P1 Latency | 28 | -0.39 | .041 |
|  | Age Adj. P1 Amplitude | 28 | -0.36 | .063 |
|  | P2 Amplitude | 28 | -0.48 | .010 |
|  | P300 Amplitude | 6 | 0.83 | .058 |
| LiSN-S Talker Advantage | Na Amplitude | 14 | -0.50 | .066 |
| NIH Picture Vocabulary Score | Early N400 Latency | 24 | -0.39 | .057 |
| NIH Dim. Card Sort Score | Pre-Wave V Latency* | 28 | -0.46 | .039 |
|  | N400 Effect Amplitude | 24 | -0.35 | .091 |
|  | Word Detect. Accuracy | 28 | 0.51 | .006 |
| NIH Total Composite Score | Pre-Wave V Latency* | 28 | -0.40 | .099 |
|  | Post-Wave V Amplitude<sup>†</sup> | 28 | -0.42 | .054 |
|  | N400 Effect Amplitude | 24 | -0.40 | .050 |
|  | Word Detect. Accuracy | 28 | 0.51 | .005 |

\* Bonferroni adjusted for correlations with Wave V and post-Wave V latencies
<sup>†</sup> Bonferroni adjusted for correlation with Wave V amplitude

**Supplementary Table 3:** Summary of all comparisons between the TD and LiD groups. Test statistics and effect sizes are reported as absolute values.

| Variable | Test Performed | Test Statistic | *df* | *p* | *d* |
| --- | --- | --- | --- | --- | --- |
| **ABR (Cz/Ch1)** |  |  |  |  |  |
| Wave V latency (ms) | Student's *t* | 1.74 | 24 | .094 | 0.68 |
| Wave V amplitude (μV) | Student's *t* | 1.73 | 24 | .097 | 0.68 |
| **LLAER (Cz/Ch1)** |  |  |  |  |  |
| Age adj. P1 latency (ms) | Student's *t* | 1.62 | 24 | .119 | 0.63 |
| Age adj. P1 amplitude (μV) | Student's *t* | 0.04 | 24 | .968 | 0.02 |
| P2 latency (ms) | Student's *t* | 0.57 | 24 | .573 | 0.22 |
| P2 amplitude (μV) | Student's *t* | 0.12 | 24 | .909 | 0.05 |
| SN latency (ms) | Wilcoxon | 90 | N = 26 | .797 | 0.13 |
| SN amplitude (μV) | Student's *t* | 0.57 | 24 | .572 | 0.22 |
| **N400 Effect** |  |  |  |  |  |
| Amplitude at Fp (μV) |  |  |  |  |  |
| Difference (Low – High) | Student's *t* | 2.08 | 18 | .052 | 0.93 |

## References

Abdollahi, F. Z., Lotfi, Y., Moosavi, A., & Bakhshi, E. (2019). Binaural interaction component of middle latency response in children suspected to central auditory processing disorder. Indian Journal of Otolaryngology and Head & Neck Surgery, 71(2), 182–185. 10.1007/s12070-017-1114-5

Allen, P., & Allan, C. (2014). Auditory processing disorders: Relationship to cognitive processes and underlying auditory neural integrity. International Journal of Pediatric Otorhinolaryngology, 78(2), 198–208. 10.1016/j.ijporl.2013.10.048

Alonso, R., & Schochat, E. (2009). The efficacy of formal auditory training in children with (central) auditory processing disorder: Behavioral and electrophysiological evaluation. 75(5), 726–732.

American Speech-Language-Hearing Association. (2005). (Central) auditory processing disorders.

Anderson, S., Skoe, E., Chandrasekaran, B., Zecker, S., & Kraus, N. (2010). Brainstem correlates of speech-in-noise perception in children. Hearing Research, 270(1–2), 151–157. 10.1016/j.heares.2010.08.001

Ankmnal-Veeranna, S., Allan, C., & Allen, P. (2019). Auditory brainstem responses in children with auditory processing disorder. Journal of the American Academy of Audiology, 30(10), 904–917. 10.3766/jaaa.18046

Backer, K. C., Kessler, A. S., Lawyer, L. A., Corina, D. P., & Miller, L. M. (2019). A novel EEG paradigm to simultaneously and rapidly assess the functioning of auditory and visual pathways. Journal of Neurophysiology, 122(4), 1312–1329. 10.1152/jn.00868.2018

Bader, R., Tarantini, L., & Mecklinger, A. (2023). Task context dissociates the FN400 and the N400. Psychophysiology, 60(7), e14258. 10.1111/psyp.14258

Barry, J. G., & Moore, D. R. (2021). *ECLiPS: Evaluation of Children’s Listening and Processing Skills* (Second). Cincinnati Children’s Hospital Medical Center.

Barry, J. G., Tomlin, D., Moore, D. R., & Dillon, H. (2015). Use of questionnaire-based measures in the assessment of listening difficulties in school-aged children. Ear and Hearing, 36(6), e300–e313.

Baum, F. L. (1901). American Fairy Tales. George M. Hill Company.

Bell, S. L., Smith, D. C., Allen, R., & Lutman, M. E. (2004). Recording the middle latency response of the auditory evoked potential as a measure of depth of anaesthesia. A technical note. British Journal of Anaesthesia, 92(3), 442–445. 10.1093/bja/aeh074

Berg, E. A. (1948). A simple objective technique for measuring flexibility in thinking. The Journal of General Psychology, 39(1), 15–22. 10.1080/00221309.1948.9918159

Best, V., Keidser, G., Buchholz, J. M., & Freeston, K. (2016). Development and preliminary evaluation of a new test of ongoing speech comprehension. International Journal of Audiology, 55(1), 45–52. 10.3109/14992027.2015.1055835

Biacabe, B., Chevallier, J. M., Avan, P., & Bonfils, P. (2001). Functional anatomy of auditory brainstem nuclei: Application to the anatomical basis of brainstem auditory evoked potentials. Auris Nasus Larynx, 28(1), 85–94. 10.1016/S0385-8146(00)00080-8

Bridger, E. K., Bader, R., Kriukova, O., Unger, K., & Mecklinger, A. (2012). The FN400 is functionally distinct from the N400. NeuroImage, 63(3), 1334–1342. 10.1016/j.neuroimage.2012.07.047

Brown, D. K., Cameron, S., Martin, J. S., Watson, C., & Dillon, H. (2010). The North American listening in spatialized noise—Sentences test (NA LiSN-S): Normative data and test-retest reliability studies for adolescents and young adults. Journal of the American Academy of Audiology, 21(10), 629–641.

Cameron, S., & Dillon, H. (2007). Development of the Listening in Spatialized Noise-Sentences Test (LISN-S). Ear and Hearing, 28(2), 196–211.

Carroll, R., Warzybok, A., Kollmeier, B., & Ruigendijk, E. (2016). Age-related differences in lexical access relate to speech recognition in noise. Frontiers in Psychology, 7. 10.3389/fpsyg.2016.00990

Čeponienė, R., Alku, P., Westerfield, M., Torki, M., & Townsend, J. (2005). ERPs differentiate syllable and nonphonetic sound processing in children and adults. Psychophysiology, 42(4), 391–406. 10.1111/j.1469-8986.2005.00305.x

Čeponienė, R., Torki, M., Alku, P., Koyama, A., & Townsend, J. (2008). Event-related potentials reflect spectral differences in speech and non-speech stimuli in children and adults. Clinical Neurophysiology, 119(7), 1560–1577. 10.1016/j.clinph.2008.03.005

Corina, D. P., Coffey-Corina, S., Pierotti, E., Bormann, B., LaMarr, T., Lawyer, L., Backer, K. C., & Miller, L. M. (2022). Electrophysiological examination of ambient speech processing in children with cochlear implants. Journal of Speech, Language, and Hearing Research, 65(9), 3502–3517. 10.1044/2022_JSLHR-22-00004

Crowley, K. E., & Colrain, I. M. (2004). A review of the evidence for P2 being an independent component process: Age, sleep and modality. Clinical Neurophysiology, 115(4), 732–744. 10.1016/j.clinph.2003.11.021

D’Arcy, R. C. N., Ghosh Hajra, S., Liu, C., Sculthorpe, L. D., & Weaver, D. F. (2011). Towards brain first-aid: A diagnostic device for conscious awareness. IEEE Transactions on Biomedical Engineering, 58(3), 750–754. 10.1109/TBME.2010.2090880

Dave, S., Brothers, T., Hoversten, L. J., Traxler, M. J., & Swaab, T. Y. (2021). Cognitive control mediates age-related changes in flexible anticipatory processing during listening comprehension. Brain Research, 1768, 147573. 10.1016/j.brainres.2021.147573

Delorme, A., & Makeig, S. (2004). EEGLAB: An open source toolbox for analysis of single-trial EEG dynamics including independent component analysis. Journal of Neuroscience Methods, 134(1), 9–21. 10.1016/j.jneumeth.2003.10.009

Denys, S., Barry, J., Moore, D. R., Verhaert, N., & Van Wieringen, A. (2024). A multi-sample comparison and Rasch analysis of the Evaluation of Children’s Listening and Processing Skills Questionnaire. Ear & Hearing, 45(5), 1202–1215. 10.1097/AUD.0000000000001509

Di Liberto, G. M., O’Sullivan, J. A., & Lalor, E. C. (2015). Low-frequency cortical entrainment to speech reflects phoneme-level processing. Current Biology, 25(19), 2457–2465. 10.1016/j.cub.2015.08.030

Dillon, H., & Cameron, S. (2021). Separating the causes of listening difficulties in children. Ear and Hearing, 42(5), 1097. 10.1097/AUD.0000000000001069

Dillon, H., Gaikwad, S., Luengtaweekul, P., Buchholz, J., & Cameron, S. (2025). Development of the Test of Listening Difficulties– Universal and Australian normative data in children and adults. Journal of Speech Language and Hearing Research, 68, 6089–6099.

Duncan, C. C., Barry, R. J., Connolly, J. F., Fischer, C., Michie, P. T., Näätänen, R., Polich, J., Reinvang, I., & Van Petten, C. (2009). Event-related potentials in clinical research: Guidelines for eliciting, recording, and quantifying mismatch negativity, P300, and N400. Clinical Neurophysiology, 120(11), 1883–1908. 10.1016/j.clinph.2009.07.045

Dwivedi, V. D., & Gibson, R. M. (2017). An ERP investigation of quantifier scope ambiguous sentences: Evidence for number in events. Journal of Neurolinguistics, 42, 63–82.

Eggermont, J. J. (2019). Chapter 30—Auditory brainstem response. In Clinical Neurophysiology: Basis and Technical Aspects (Vol. 160, pp. 451–464). Elsevier.

Falcone, H., Denys, S., Verhaert, N., & Van Wieringen, A. (2026). Using behavioral tasks to probe listening difficulties in normal hearing or near-to-normal hearing children up to 14 years old: A systematic review. Ear & Hearing, 47(3), 585–595. 10.1097/AUD.0000000000001758

Ferguson, M. A., Hall, R. L., Riley, A., & Moore, D. R. (2011). Communication, listening, cognitive and speech perception skills in children with Auditory Processing Disorder (APD) or Specific Language Impairment (SLI). Journal of Speech, Language, and Hearing Research, 54(1), 211–227. 10.1044/1092-4388(2010/09-0167)

Fields, R. D. (2008). White matter in learning, cognition and psychiatric disorders. Trends in Neurosciences, 31(7), 361–370. 10.1016/j.tins.2008.04.001

Filippini, R., & Schochat, E. (2009). Brainstem evoked auditory potentials with speech stimulus in the auditory processing disorder. Brazilian Journal of Otorhinolaryngology, 75(3), 449–455. 10.1016/S1808-8694(15)30665-0

Fox, J., & Weisberg, S. (2019). An {R} Companion to Applied Regression (3rd Edition). Sage. https://socialsciences.mcmaster.ca/jfox/Books/Companion/

Gopal, K. V., Daily, C. S., & Kao, K. (2002). Auditory brainstem responses to regular and high stimulus repetition rates in children at risk for central auditory processing disorders. Journal of Audiological Medicine, 11(3), 146–160.

Grunwald, T., Boutros, N. N., Pezer, N., Von Oertzen, J., Fernández, G., Schaller, C., & Elger, C. E. (2003). Neuronal substrates of sensory gating within the human brain. Biological Psychiatry, 53(6), 511–519. 10.1016/S0006-3223(02)01673-6

Hajimohammadi, A., Bagheri, S., Fatemi, N., Amanollahi, A., & Rasouli Fard, P. (2025). Applications of P300 in identifying and monitoring auditory processing disorders: A systematic review and meta-analysis. *Speech*, Language and Hearing, 28(1), 2592406. 10.1080/2050571X.2025.2592406

Hamza, Y., Yang, Y., Vu, J., Abdelmalek, A., Malekifar, M., Barnes, C. A., & Zeng, F.-G. (2024). Auditory brainstem responses as a biomarker for cognition. Communications Biology, 7(1), 1653. 10.1038/s42003-024-07346-4

Hind, S. E., Haines-Bazrafshan, R., Benton, C. L., Brassington, W., Towle, B., & Moore, D. R. (2011). Prevalence of clinical referrals having hearing thresholds within normal limits. International Journal of Audiology, 50(10), 708–716.

Humanski, R. A., & Butler, R. A. (1988). The contribution of the near and far ear toward localization of sound in the sagittal plane. The Journal of the Acoustical Society of America, 83(6), 2300–2310.

Hunter, L. L., Blankenship, C. M., Lin, L., Sloat, N. T., Perdew, A., Stewart, H., & Moore, D. R. (2020). Peripheral auditory involvement in childhood listening difficulty. Ear and Hearing, 42(1), 29–41. doi:%2010.1097/AUD.0000000000000899

Hunter, L. L., Blankenship, C. M., Shinn-Cunningham, B., Hood, L., Motlagh Zadeh, L., & Moore, D. R. (2023). Brainstem auditory physiology in children with listening difficulties. Hearing Research, 429, 108705.

Jamison, C., Aiken, S. J., Kiefte, M., Newman, A. J., Bance, M., & Sculthorpe-Petley, L. (2016). Preliminary investigation of the passively evoked N400 as a tool for estimating speech-in-noise thresholds. American Journal of Audiology, 25(4), 344–358. 10.1044/2016_AJA-15-0080

Jeon, Y., & Polich, J. (2003). Meta analysis of P300 and schizophrenia: Patients, paradigms, and practical implications. Psychophysiology, 40(5), 684–701. 10.1111/1469-8986.00070

Jerger, J., & Musiek, F. (2000). Report of the consensus conference on the diagnosis of auditory processing disorders in school-aged children. Journal of the American Academy of Audiology, 11(9), 8.

Jirsa, R. E. (1992). The clinical utility of the P3 AERP in children with auditory processing disorders. Journal of Speech, Language, and Hearing Research, 35(4), 903–912. 10.1044/jshr.3504.903

Jirsa, R. E. (2001). Maximum length sequences-auditory brainstem responses from children with auditory processing disorders. Journal of the American Academy of Audiology, 12(03), 155–164. 10.1055/s-0042-1745592

Jirsa, R. E., & Clontz, K. B. (1990). Long latency auditory event-related potentials from children with auditory processing disorders. Ear and Hearing, 11(3), 222–232.

Johnson, K. L., Nicol, T. G., & Kraus, N. (2005). Brain stem response to speech: A biological marker of auditory processing. Ear & Hearing, 26(5), 424–434. 10.1097/01.aud.0000179687.71662.6e

Joint Committee on Infant Hearing. (2007). Year 2007 position statement: Principles and guidelines for early hearing detection and intervention programs. Pediatrics, (120), 898–921.

Joos, K., Gilles, A., Van De Heyning, P., De Ridder, D., & Vanneste, S. (2014). From sensation to percept: The neural signature of auditory event-related potentials. Neuroscience & Biobehavioral Reviews, 42, 148–156. 10.1016/j.neubiorev.2014.02.009

Kaandorp, M. W., De Groot, A. M. B., Festen, J. M., Smits, C., & Goverts, S. T. (2016). The influence of lexical-access ability and vocabulary knowledge on measures of speech recognition in noise. International Journal of Audiology, 55(3), 157–167.

Kallioinen, P., Olofsson, J., Nakeva Von Mentzer, C., Lindgren, M., Ors, M., Sahlén, B. S., Lyxell, B., Engström, E., & Uhlén, I. (2016). Semantic processing in deaf and hard-of-hearing children: Large N400 mismatch effects in brain responses, despite poor semantic ability. Frontiers in Psychology, 7. 10.3389/fpsyg.2016.01146

Kassambara, A. (2020). rstatix: Pipe-Friendly Framework for Basic Statistical Tests (Version 0.6.0) [Computer software]. https://CRAN.R-project.org/package=rstatix

Klug, M., & Gramann, K. (2021). Identifying key factors for improving ICA-based decomposition of EEG data in mobile and stationary experiments. European Journal of Neuroscience, 54(12), 8406–8420.

Klumpp, H., Keller, J., Miller, G. A., Casas, B. R., Best, J. L., & Deldin, P. J. (2010). Semantic processing of emotional words in depression and schizophrenia. International Journal of Psychophysiology, 75(2), 211–215. 10.1016/j.ijpsycho.2009.12.004

Kojima, K., Lin, L., Petley, L., Clevenger, N., Perdew, A., Bodik, M., Blankenship, C. M., Motlagh Zadeh, L., Hunter, L. L., & Moore, D. R. (2024). Childhood listening and associated cognitive difficulties persist into adolescence. Ear and Hearing, 45(5), 1252–1263. 10.1097/AUD.0000000000001517

Koravand, A., Jutras, B., & Lassonde, M. (2017). Abnormalities in cortical auditory responses in children with central auditory processing disorder. Neuroscience, 346, 135–148. 10.1016/j.neuroscience.2017.01.011

Krishnamurti, S. (2001). P300 auditory event-related potentials in binaural and competing noise conditions in adults with central auditory processing disorders. Contemporary Issues in Communication Science and Disorders, 28(Spring), 40–47. 10.1044/cicsd_28_S_40

Kutas, M., & Federmeier, K. D. (2011). Thirty years and counting: Finding meaning in the N400 component of the event-related brain potential (ERP). Annual Review of Psychology, 62(1), 621– 647. 10.1146/annurev.psych.093008.131123

Kutas, M., & Hillyard, S. A. (1984). Brain potentials during reading reflect word expectancy and semantic association. Nature, 307(5947), 161–163.

Lau, E. F., Phillips, C., & Poeppel, D. (2008). A cortical network for semantics: (De)constructing the N400. Nature Reviews Neuroscience, 9(12), 920–933. 10.1038/nrn2532

Legatt, A. D., Arezzo, J. C., & Vaughan, H. G. (1988). The anatomic and physiologic bases of brain stem auditory evoked potentials. Neurologic Clinics, 6(4), 681–704.

Liasis, A., Bamiou, D.-E., Campbell, P., Sirimanna, T., Boyd, S., & Towell, A. (2003). Auditory Event-Related Potentials in the Assessment of Auditory Processing Disorders: A Pilot Study. Neuropediatrics, 34(1), 23–29. 10.1055/s-2003-38622

Lopez-Calderon, J., & Luck, S. J. (2014). ERPLAB: An open-source toolbox for the analysis of event-related potentials. Frontiers in Human Neuroscience, 8, 213. 10.3389/fnhum.2014.00213

Magimairaj, B. M., Nagaraj, N. K., Sergeev, A. V., & Benafield, N. J. (2020). Comparison of auditory, language, memory, and attention abilities in children with and without listening difficulties. American Journal of Audiology, 29(4), 710–727. 10.1044/2020_AJA-20-00018

Mankel, K., Comstock, D. C., Bormann, B. M., Das, S., Sagiv, D., Brodie, H., & Miller, L. M. (2024). Auditory brainstem responses to speech-in-noise reflect selective attention, comprehension, and subjective listening effort. Neuroscience. 10.1101/2024.12.23.629710

Mankel, K., Comstock, D. C., Bormann, B. M., Das, S., Sagiv, D., Brodie, H., & Miller, L. M. (in press). Auditory brainstem encoding of speech-in-noise predicts word identification, narrative comprehension, and subjective listening effort in a selective attention task. eNeuro.

Marchand, Y., D’Arcy, R. C. N., & Connolly, J. F. (2002). Linking neurophysiological and neuropsychological measures for aphasia assessment. Clinical Neurophysiology, 113(11), 1715– 1722. 10.1016/S1388-2457(02)00224-9

Mattsson, T. S., Lind, O., Follestad, T., Grøndahl, K., Wilson, W., Nicholas, J., Nordgård, S., & Andersson, S. (2019). Electrophysiological characteristics in children with listening difficulties, with or without auditory processing disorder. International Journal of Audiology, 58(11), 704– 716. 10.1080/14992027.2019.1621396

McGrath, M. A., Fletcher, K. L., & Bielski, L. M. (2023). Executive functioning skills of children with listening difficulties. Psychology in the Schools, pits.22940. 10.1002/pits.22940

Melcher, J. R., & Kiang, N. Y. S. (1996). Generators of the brainstem auditory evoked potential in cat III: Identified cell populations. Hearing Research, 93(1–2), 52–71. 10.1016/0378-5955(95)00200-6

Miller, G. A., Heise, G. A., & Lichten, W. (1951). The intelligibility of speech as a function of the context of the test materials. Journal of Experimental Psychology, 41(5), 329–335. 10.1037/h0062491

Miller, G. A., & Isard, S. (1963). Some perceptual consequences of linguistic rules. Journal of Verbal Learning and Verbal Behavior, 2(3), 217–228. 10.1016/S0022-5371(63)80087-0

Miller, L. M., & Moore IV, B. (2020). Frequency-multiplexed speech-sound stimuli for hierarchical neural characterization of speech processing. (Patent No. 10729387).

Moore, D. R. (2006). Auditory processing disorder (APD): Definition, diagnosis, neural basis, and intervention. Audiological Medicine, 4(1), 4–11. 10.1080/16513860600568573

Moore, D. R., & Dillon, H. (2018). How should we detect and identify deficit specific auditory processing disorders. ENT & Audiology News, 27, 73–74.

Moore, D. R., Ferguson, M. A., Edmondson-Jones, A. M., Ratib, S., & Riley, A. (2010). Nature of auditory processing disorder in children. PEDIATRICS, 126(2), e382–e390.

Moore, D. R., Hugdahl, K., Stewart, H. J., Vannest, J., Perdew, A. J., Sloat, N. T., Cash, E. K., & Hunter, L. L. (2020). Listening difficulties in children: Behavior and brain activation produced by dichotic listening of CV syllables. Frontiers in Psychology, 11, 675. 10.3389/fpsyg.2020.00675

Moore, D. R., Sieswerda, S. L., Grainger, M. M., Bowling, A., Smith, N., Perdew, A., Eichert, S., Alston, S., Hilbert, L. W., Summers, L., Lin, L., & Hunter, L. L. (2018). Referral and diagnosis of developmental Auditory Processing Disorder in a large, United States hospital-based audiology service. Journal of the American Academy of Audiology, 29(5), 364–377. 10.3766/jaaa.16130

Morlet, T., Nagao, K., Greenwood, L. A., Cardinale, R. M., Gaffney, R. G., & Riegner, T. (2019). Auditory event-related potentials and function of the medial olivocochlear efferent system in children with auditory processing disorders. International Journal of Audiology, 58(4), 213–223. 10.1080/14992027.2018.1551632

Musiek, F., Gollegly, K., Lamb, L., & Lamb, P. (1990). Selected issues in screening for central auditory processing dysfunction. Seminars in Hearing, 11(04), 372–383. 10.1055/s-0028-1085516

Musiek, F., & Nagle, S. (2018). The middle latency response: A review of findings in various central nervous system lesions. Journal of the American Academy of Audiology, 29(09), 855–867. 10.3766/jaaa.16141

Näätänen, R., & Picton, T. W. (1987). The N1 wave of the human electric and magnetic response to sound: A review and an analysis of the component structure. Psychophysiology, 24(4), 375–425.

Olichney, J. M., Taylor, J. R., Gatherwright, J., Salmon, D. P., Bressler, A. J., Kutas, M., & Iragui-Madoz, V. J. (2008). Patients with MCI and N400 or P600 abnormalities are at very high risk for conversion to dementia. Neurology, 70(19_part_2), 1763–1770. 10.1212/01.wnl.0000281689.28759.ab

Omidvar, S., Duquette-Laplante, F., Bursch, C., Jutras, B., & Koravand, A. (2023). Assessing auditory processing in children with listening difficulties: A pilot study. Journal of Clinical Medicine, 12(3), 897. 10.3390/jcm12030897

Pascoinelli, A. T., Schochat, E., & Murphy, C. F. B. (2021). Executive function and sensory processing in dichotic listening of young adults with listening difficulties. Journal of Clinical Medicine, 10(18), 4255. 10.3390/jcm10184255

Petley, L., Bardouille, T., Chiasson, D., Froese, P., Patterson, S., Newman, A., Omisade, A., & Beyea, S. (2018). Attentional dysfunction and recovery in concussion: Effects on the P300m and contingent magnetic variation. Brain Injury, 32(4), 464–473. 10.1080/02699052.2018.1429022

Petley, L., Blankenship, C., Hunter, L. L., Stewart, H. J., Lin, L., & Moore, D. R. (2024). Amplitude modulation perception and cortical evoked potentials in children with listening difficulties and their typically developing peers. Journal of Speech, Language, and Hearing Research, 67, 633– 656.

Petley, L., Hunter, L. L., Zadeh, L. M., Stewart, H. J., Sloat, N. T., Perdew, A., Lin, L., & Moore, D. R. (2021). Listening difficulties in children with normal audiograms: Relation to hearing and cognition. Ear and Hearing, 42(6), 1640.

Pichora-Fuller, M. K., Kramer, S. E., Eckert, M. A., Edwards, B., Hornsby, B. W. Y., Humes, L. E., Lemke, U., Lunner, T., Matthen, M., Mackersie, C. L., Naylor, G., Phillips, N. A., Richter, M., Rudner, M., Sommers, M. S., Tremblay, K. L., & Wingfield, A. (2016). Hearing impairment and cognitive energy: The Framework for Understanding Effortful Listening (FUEL). Ear and Hearing, 37(1), 5S–27S. 10.1097/AUD.0000000000000312

Picton, T. W. (2010). Human auditory evoked potentials. Plural Publishing.

Pijnacker, J., Davids, N., Van Weerdenburg, M., Verhoeven, L., Knoors, H., & Van Alphen, P. (2017). Semantic processing of sentences in preschoolers with Specific Language Impairment: Evidence from the N400 effect. Journal of Speech, Language, and Hearing Research, 60(3), 627–639. 10.1044/2016_JSLHR-L-15-0299

Pillion, J. P., Boatman-Reich, D., & Gordon, B. (2018). Auditory brainstem pathology in Autism Spectrum Disorder: A review. Cognitive and Behavioral Neurology, 31(2), 53–78. 10.1097/WNN.0000000000000154

Polich, J. (2007). Updating P300: An integrative theory of P3a and P3b. Clinical Neurophysiology, 118(10), 2128–2148. 10.1016/j.clinph.2007.04.019

Ponton, C., Eggermont, J. J., Khosla, D., Kwong, B., & Don, M. (2002). Maturation of human central auditory system activity: Separating auditory evoked potentials by dipole source modeling. Clinical Neurophysiology, 113(3), 407–420. 10.1016/S1388-2457(01)00733-7

Ponton, C. W., Moore, J. K., & Eggermont, J. J. (1996). Auditory brain stem response generation by parallel pathways: Differential maturation of axon conduction time and synaptic transmission. Ear & Hearing, 17(5), 402–410.

Purdy, S. C., Kelly, A. S., & Davies, M. G. (2002). Auditory brainstem response, middle latency response, and late cortical evoked potentials in children with learning disabilities. Journal of the American Academy of Audiology, 13(07), 367–382. 10.1055/s-0040-1715981

Rocha-Muniz, C. N., Befi-Lopes, D. M., & Schochat, E. (2012). Investigation of auditory processing disorder and language impairment using the speech-evoked auditory brainstem response. Hearing Research, 294(1–2), 143–152. 10.1016/j.heares.2012.08.008

Schochat, E., Musiek, F. E., Alonso, R., & Ogata, J. (2010). Effect of auditory training on the middle latency response in children with (central) auditory processing disorder. Brazilian Journal of Medical and Biological Research, 43(8), 777–785. 10.1590/S0100-879X2010007500069

Sculthorpe-Petley, L., Liu, C., Ghosh Hajra, S., Parvar, H., Satel, J., Trappenberg, T. P., Boshra, R., & D’Arcy, R. C. N. (2015). A rapid event-related potential (ERP) method for point-of-care evaluation of brain function: Development of the Halifax Consciousness Scanner. Journal of Neuroscience Methods, 245, 64–72. 10.1016/j.jneumeth.2015.02.008

Selvanayagam, J., Witte, V., Schmidt, L. A., & Dwivedi, V. D. (2019). A preliminary investigation of dispositional affect, the P300, and sentence processing. Brain Research, 1721, 146309.

Sharma, A., Kraus, N., J. McGee, T., & Nicol, T. G. (1997). Developmental changes in P1 and N1 central auditory responses elicited by consonant-vowel syllables. Electroencephalography and Clinical Neurophysiology/Evoked Potentials Section, 104(6), 540–545. 10.1016/S0168-5597(97)00050-6

Sharma, M., Dhamani, I., Leung, J., & Carlile, S. (2014). Attention, memory, and auditory processing in 10-to 15-year-old children with listening difficulties. *Journal of Speech*, Language, and Hearing Research, 57(6), 2308–2321. 10.1044/2014_JSLHR-H-13-0226

Sharma, M., Purdy, S. C., & Humburg, P. (2019). Cluster analyses reveals subgroups of children With suspected auditory processing disorders. Frontiers in Psychology, 10, 2481. 10.3389/fpsyg.2019.02481

Shehabi, S., Comstock, D. C., Mankel, K., Bormann, B. M., Das, S., Brodie, H., Sagiv, D., & Miller, L. M. (2025). Individual differences in cognition and perception predict neural processing of speech in noise for audiometrically normal listeners. eNeuro, 12(4), ENEURO.0381-24.2025. 10.1523/ENEURO.0381-24.2025

Song, J. H., Banai, K., Russo, N. M., & Kraus, N. (2006). On the relationship between speech-and nonspeech-evoked auditory brainstem responses. Audiology and Neurotology, 11(4), 233–241. 10.1159/000093058

Spearman, C. (1904). “General Intelligence,” objectively determined and measured. The American Journal of Psychology, 15, 201–292.

Stewart, H. J., Cash, E. K., Hunter, L. L., Maloney, T., Vannest, J., & Moore, D. R. (2022). Speech cortical activation and connectivity in typically developing children and those with listening difficulties. NeuroImage: Clinical, 36, 103172. 10.1016/j.nicl.2022.103172

Strauss, D. J., Corona-Strauss, F. I., Trenado, C., Bernarding, C., Reith, W., Latzel, M., & Froehlich, M. (2010). Electrophysiological correlates of listening effort: Neurodynamical modeling and measurement. Cognitive Neurodynamics, 4(2), 119–131. 10.1007/s11571-010-9111-3

Taylor, W. L. (1953). “Cloze Procedure”: A new tool for measuring readability. Journalism Quarterly, 30(4), 415–433. 10.1177/107769905303000401

Tomlin, D., & Rance, G. (2016). Maturation of the central auditory nervous system in children with auditory processing disorder. Seminars in Hearing, 37(01), 074–083. 10.1055/s-0035-1570328

Torchiano, M. (2020). effsize: Efficient effect size computation (Version 0.8.1) [Computer software]. 10.5281/zenodo.1480624

Voss, J. L., & Federmeier, K. D. (2011). FN400 potentials are functionally identical to N400 potentials and reflect semantic processing during recognition testing. Psychophysiology, 48(4), 532–546. 10.1111/j.1469-8986.2010.01085.x

Walker, E. A., Sapp, C., Oleson, J. J., & McCreery, R. W. (2019). Longitudinal speech recognition in noise in children: Effects of hearing status and vocabulary. Frontiers in Psychology, 10, 2421. 10.3389/fpsyg.2019.02421

Weintraub, S., Dikmen, S. S., Heaton, R. K., Tulsky, D. S., Zelazo, P. D., Bauer, P. J., Carlozzi, N. E., Slotkin, J., Blitz, D., Wallner-Allen, K., Fox, N. A., Beaumont, J. L., Mungas, D., Nowinski, C. J., Richler, J., Deocampo, J. A., Anderson, J. E., Manly, J. J., Borosh, B.,…Gershon, R. C. (2013). Cognition assessment using the NIH Toolbox. Neurology, 80(Issue 11, Supplement 3), S54–S64.

Wilson, W. J., & Arnott, W. (2013). Using different criteria to diagnose (central) Auditory Processing Disorder: How big a difference does it make? Journal of Speech, Language, and Hearing Research, 56(1), 63–70. 10.1044/1092-4388(2012/11-0352)

Winkler, I., Debener, S., Muller, K.-R., & Tangermann, M. (2015). On the influence of high-pass filtering on ICA-based artifact reduction in EEG-ERP. 2015 37th Annual International Conference of the IEEE Engineering in Medicine and Biology Society (EMBC), 4101–4105. 10.1109/EMBC.2015.7319296

Zhou, X., Dillon, H., Tomlin, D., Burgoyne, K., Gurteen, H., Nixon, G., Gudkar, A. I., & Heinrich, A. (2026). Assessing language skills as a predictor of children’s listening difficulties: Validation and reference data on a new auditory language task. Ear & Hearing, 47(1), 226–235. 10.1097/AUD.0000000000001715

Zion Golumbic, E. M., Ding, N., Bickel, S., Lakatos, P., Schevon, C. A., McKhann, G. M., Goodman, R. R., Emerson, R., Mehta, A. D., Simon, J. Z., Poeppel, D., & Schroeder, C. E. (2013). Mechanisms underlying selective neuronal tracking of attended speech at a “cocktail party.” Neuron, 77(5), 980–991. 10.1016/j.neuron.2012.12.037

